# Linafexor, a Pulsatile FXR Agonist, Augments Low-Dose Semaglutide for Weight Loss in Adults with Obesity, MASH, and Type 2 Diabetes

**DOI:** 10.64898/2026.10.18.26365821

**Authors:** Yifei Zhang, Yufei Chen, Shan Huang, Lei Jiang, Yufan Wang, Jing Ma, Rong Deng, Juan Du, Lili Xia, Yuhang Ma, Peihong Wu, Weituo Zhang, Cong Liu, Guanguan Zhao, Gaihong Wang, Mengyi Qian, Huimin Lin, Fuhua Yan, Guang Ning, H. Eric Xu, Weiqing Wang

## Abstract

Glucagon-like peptide-1 (GLP-1) receptor agonists produce less weight loss in people with type 2 diabetes than without, and higher compensatory doses are limited by gastrointestinal adverse events and lean-mass loss—leaving patients with obesity, type 2 diabetes, and metabolic dysfunction-associated steatohepatitis (MASH) inadequately treated. We tested whether linafexor, a pulsatile farnesoid X receptor (FXR) agonist, could augment the weight-loss efficacy of low-dose semaglutide in this population while preserving tolerability. In this multicenter, randomized, double-blind, placebo-controlled, phase 2 trial, 62 adults with obesity (body-mass index, 30– 45 kg/m²), treatment-naive type 2 diabetes, and MASH (MRI-PDFF ≥10%) were assigned to linafexor (4 mg orally once daily) or placebo, each with semaglutide (0.5 mg once weekly), for 16 weeks. The primary outcome was the percentage change in body weight at week 16. Adding linafexor more than doubled weight loss (−6.17% vs. −2.91%; treatment difference, −3.26%; 95% CI, −5.35 to −1.18; P=0.003), and more participants achieved at least 5% weight loss (53.3% vs. 28.1%; P=0.046). Consistent with this, combination therapy further reduced visceral adipose tissue (treatment difference, −13.85 cm²; P=0.035), the liver enzymes alanine aminotransferase and γ-glutamyl transferase, and serum bile acids, whose changes correlated with weight and liver-enzyme improvements—associations absent with monotherapy. Gastrointestinal adverse events were comparable (23.3% vs. 18.8%), with no treatment-related serious adverse events or discontinuations. Thus, in a population with blunted GLP-1 responses, adding linafexor to low-dose semaglutide enhanced weight loss and improved body composition and hepatic measures while preserving tolerability—a dose-sparing, mechanism-complementary strategy (ClinicalTrials.gov number, NCT06492330).

## Introduction

Obesity and its metabolic sequelae—type 2 diabetes and metabolic dysfunction-associated steatohepatitis (MASH)—frequently coexist and together constitute a profound and growing global health burden^1–3^. Glucagon-like peptide-1 (GLP-1) receptor agonists such as semaglutide have transformed obesity management, producing substantial weight loss at high doses^4–7^. Their real-world utility, however, is constrained by dose-dependent gastrointestinal adverse events that drive early discontinuation^4,8^, so that many patients never reach the doses required for maximal benefit^9,10^. High-dose therapy also produces a disproportionate loss of lean muscle mass^11,12^. Most pertinent to the patients considered here, the weight-loss response to GLP-1 receptor agonists is attenuated in people with type 2 diabetes relative to those without^4,13^—precisely the population in whom obesity and MASH most often cluster. Strategies that preserve efficacy in these patients while improving tolerability are therefore urgently needed.

Combining established agents at lower doses to exploit complementary mechanisms is a rational way to maximize metabolic benefit while limiting dose-related toxicity. The farnesoid X receptor (FXR), a bile acid–activated nuclear receptor, is an attractive partner for this purpose: it governs bile acid synthesis, lipid and glucose homeostasis, and energy expenditure^14–17^, and is a validated therapeutic target in MASH^18^. Preclinical studies show that FXR agonism potentiates the metabolic effects of GLP-1 receptor agonists in models of obesity and MASH^19^, and FXR activation independently increases energy expenditure and promotes browning of white adipose tissue through upregulation of uncoupling protein 1 (UCP1)^20,21^. Because these actions—raising thermogenesis and improving insulin sensitivity—are distinct from the appetite suppression and delayed gastric emptying that mediate GLP-1 effects, FXR agonism could augment GLP-1–induced weight loss, counter the blunted response seen in diabetic patients, and favor preservation of lean mass.

Realizing this potential in the clinic has proved difficult. Conventional FXR agonists were designed for sustained receptor occupancy, which disrupts the normally pulsatile rhythm of bile acid signaling and produces hepatotoxicity and limited efficacy at tolerable doses^22,23^; these liabilities underlie the clinical failure or market withdrawal of more than 20 FXR-targeting candidates^23–27^. Linafexor (CS0159) was engineered to overcome this constraint: its short half-life (<1 hour) produces brief, pulsatile FXR activation aligned with physiological bile acid rhythms. In a related phase 1 program, linafexor showed an excellent safety profile with no treatment-related adverse events and effective, transient FXR pathway engagement^28^. We therefore conducted a randomized, double-blind, placebo-controlled, phase 2 trial to determine whether adding linafexor to low-dose semaglutide could enhance weight loss while maintaining tolerability in adults with obesity, type 2 diabetes, and MASH—to our knowledge one of the first clinical tests of whether pulsatile FXR agonism can augment GLP-1–based therapy in humans. We hypothesized that the combination would produce superior weight loss with tolerability comparable to that of low-dose semaglutide alone.

## Results

### Trial design and study participants

In this multicenter, randomized, double-blind, placebo-controlled phase 2 trial conducted at four sites in China, adults with obesity, type 2 diabetes, and MASH were randomly assigned in a 1:1 ratio to oral linafexor (4 mg once daily) or matching placebo, each administered with low-dose subcutaneous semaglutide (0.5 mg once weekly), for 16 weeks; randomization followed a 2-week screening period and was succeeded by a 4-week off-treatment follow-up (**Extended Data Fig. 1**; full eligibility criteria and procedures are provided in Methods).

Between July 19 and September 27, 2024, 100 participants were screened, of whom 38 did not meet eligibility criteria—most commonly because glycated hemoglobin (HbA1c; n=12) or fasting plasma glucose (n=8) fell outside the specified ranges (**Fig. 1**). The remaining 62 participants, all with MASH confirmed by a magnetic resonance imaging proton density fat fraction (MRI-PDFF) ≥10%, underwent randomization: 30 to linafexor plus semaglutide and 32 to placebo plus semaglutide, and all were included in the primary analysis. One participant in each group was lost to follow-up at week 16, so that 29 participants (96.7%) in the linafexor plus semaglutide group and 31 (96.9%) in the placebo plus semaglutide group completed the 16-week treatment period. No participant required a semaglutide dose adjustment during the trial.

**Fig 1.**
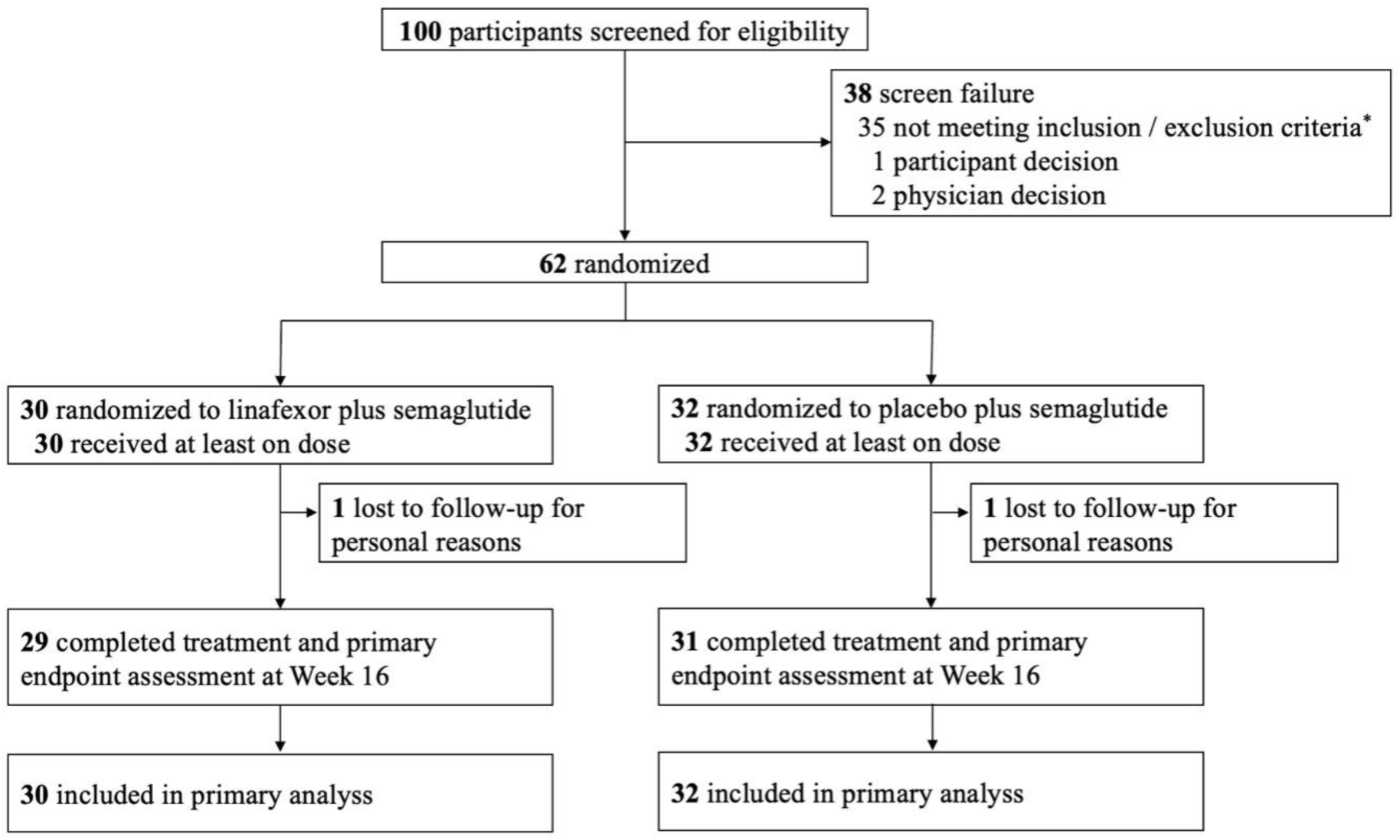
Flowchart of Trial Participants. * The most common reasons for not meeting inclusion/exclusion criteria included HbA1c outside 7.0-10.5% range (n=12), fasting plasma glucose outside 126-240 mg/dl range (n=8), and metabolic dysfunction-associated steatohepatitis not confirmed by biopsy or magnetic resonance imaging proton density fat fraction (n=5).

Baseline characteristics were well balanced between the groups (**Table 1**). The cohort was predominantly male (69%), with a mean age of 37.3 years and substantial adiposity: mean body weight 100.2 kg, mean body mass index (BMI) 34.3 kg/m², mean waist circumference 109.9 cm, and mean fat mass 38.0 kg (38.1% body fat) alongside a mean skeletal muscle mass of 34.9 kg. Metabolic disease was advanced, with a mean HbA1c of 8.6% and a mean liver fat content of 21.0% by MRI-PDFF.

**Table 1.**
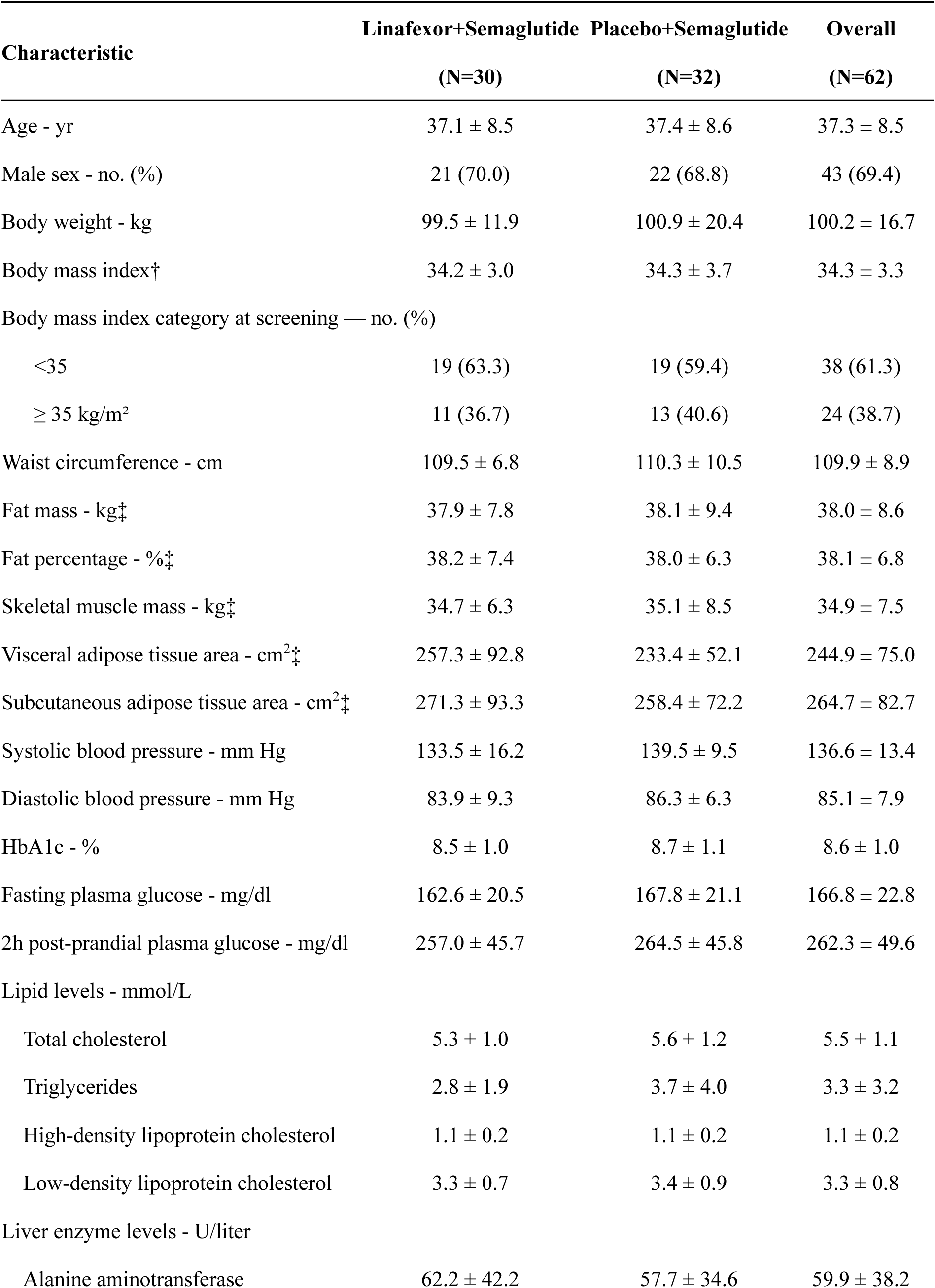
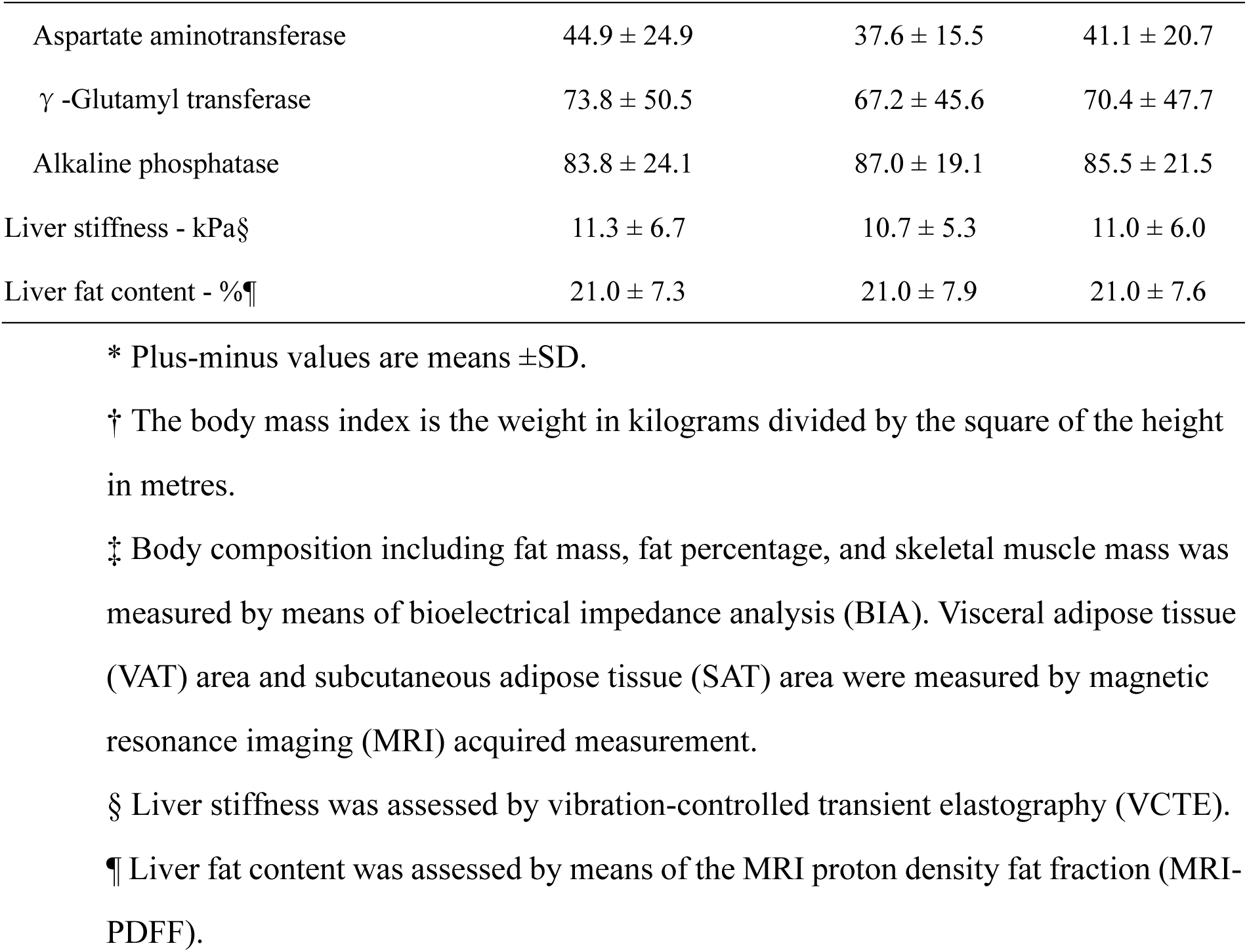
Demographic and Clinical Characteristics of the Participants at Baseline*.

| Characteristic | Linafexor+Semaglutide<br>(N=30) | Placebo+Semaglutide<br>(N=32) | Overall<br>(N=62) |
| --- | --- | --- | --- |
| Age - yr | 37.1 ± 8.5 | 37.4 ± 8.6 | 37.3 ± 8.5 |
| Male sex - no. (%) | 21 (70.0) | 22 (68.8) | 43 (69.4) |
| Body weight - kg | 99.5 ± 11.9 | 100.9 ± 20.4 | 100.2 ± 16.7 |
| Body mass index† | 34.2 ± 3.0 | 34.3 ± 3.7 | 34.3 ± 3.3 |
| Body mass index category at screening — no. (%) |  |  |  |
| <35 | 19 (63.3) | 19 (59.4) | 38 (61.3) |
| ≥ 35 kg/m<sup>2</sup> | 11 (36.7) | 13 (40.6) | 24 (38.7) |
| Waist circumference - cm | 109.5 ± 6.8 | 110.3 ± 10.5 | 109.9 ± 8.9 |
| Fat mass - kg‡ | 37.9 ± 7.8 | 38.1 ± 9.4 | 38.0 ± 8.6 |
| Fat percentage - %‡ | 38.2 ± 7.4 | 38.0 ± 6.3 | 38.1 ± 6.8 |
| Skeletal muscle mass - kg‡ | 34.7 ± 6.3 | 35.1 ± 8.5 | 34.9 ± 7.5 |
| Visceral adipose tissue area - cm<sup>2</sup>‡ | 257.3 ± 92.8 | 233.4 ± 52.1 | 244.9 ± 75.0 |
| Subcutaneous adipose tissue area - cm<sup>2</sup>‡ | 271.3 ± 93.3 | 258.4 ± 72.2 | 264.7 ± 82.7 |
| Systolic blood pressure - mm Hg | 133.5 ± 16.2 | 139.5 ± 9.5 | 136.6 ± 13.4 |
| Diastolic blood pressure - mm Hg | 83.9 ± 9.3 | 86.3 ± 6.3 | 85.1 ± 7.9 |
| HbA1c - % | 8.5 ± 1.0 | 8.7 ± 1.1 | 8.6 ± 1.0 |
| Fasting plasma glucose - mg/dl | 162.6 ± 20.5 | 167.8 ± 21.1 | 166.8 ± 22.8 |
| 2h post-prandial plasma glucose - mg/dl | 257.0 ± 45.7 | 264.5 ± 45.8 | 262.3 ± 49.6 |
| Lipid levels - mmol/L |  |  |  |
| Total cholesterol | 5.3 ± 1.0 | 5.6 ± 1.2 | 5.5 ± 1.1 |
| Triglycerides | 2.8 ± 1.9 | 3.7 ± 4.0 | 3.3 ± 3.2 |
| High-density lipoprotein cholesterol | 1.1 ± 0.2 | 1.1 ± 0.2 | 1.1 ± 0.2 |
| Low-density lipoprotein cholesterol | 3.3 ± 0.7 | 3.4 ± 0.9 | 3.3 ± 0.8 |
| Liver enzyme levels - U/liter |  |  |  |
| Alanine aminotransferase | 62.2 ± 42.2 | 57.7 ± 34.6 | 59.9 ± 38.2 |
| Aspartate aminotransferase | 44.9 ± 24.9 | 37.6 ± 15.5 | 41.1 ± 20.7 |
| γ -Glutamyl transferase | 73.8 ± 50.5 | 67.2 ± 45.6 | 70.4 ± 47.7 |
| Alkaline phosphatase | 83.8 ± 24.1 | 87.0 ± 19.1 | 85.5 ± 21.5 |
| Liver stiffness - kPa§ | 11.3 ± 6.7 | 10.7 ± 5.3 | 11.0 ± 6.0 |
| Liver fat content - %¶ | 21.0 ± 7.3 | 21.0 ± 7.9 | 21.0 ± 7.6 |

\* Plus-minus values are means ±SD.
† The body mass index is the weight in kilograms divided by the square of the height in metres.
‡ Body composition including fat mass, fat percentage, and skeletal muscle mass was measured by means of bioelectrical impedance analysis (BIA). Visceral adipose tissue (VAT) area and subcutaneous adipose tissue (SAT) area were measured by magnetic resonance imaging (MRI) acquired measurement.
§ Liver stiffness was assessed by vibration-controlled transient elastography (VCTE).
¶ Liver fat content was assessed by means of the MRI proton density fat fraction (MRI-PDFF).

### Change in body weight

Adding linafexor to low-dose semaglutide more than doubled weight loss. In the trial product estimand analysis (the primary estimand), the mean percentage change in body weight from baseline to week 16 was −6.17% (95% confidence interval [CI], −7.64 to −4.70) in the linafexor plus semaglutide group versus −2.91% (95% CI, −4.48 to −1.33) in the placebo plus semaglutide group (**Table 2 and Fig. 2A**), corresponding to a mixed-effects model treatment difference of −3.26% (95% CI, −5.35 to −1.18; P=0.003). The groups separated as early as week 4 and diverged progressively thereafter (**Fig. 2A and Fig. 2B**), and weight loss remained linear, without a plateau, through week 16 in both groups. The effect was consistent at the individual level: 90% of participants receiving combination therapy lost more weight than the median participant in the control group (**Fig. 2C**). Clinically meaningful responses were correspondingly more frequent—weight loss of at least 5% occurred in 53.3% of the linafexor plus semaglutide group versus 28.1% of the placebo plus semaglutide group (odds ratio, 2.92; 95% CI, 1.02 to 8.37; P=0.046), and weight loss of at least 10% in 16.7% versus 6.3%, respectively (**Fig. 2D**). The benefit was consistent across prespecified subgroups, with no significant interaction effects (**Extended Data Fig. 2**). Results for the treatment policy estimand and the per-protocol set were consistent with the primary analysis and are provided in **Extended Data Tables 1 and 2**.

**Fig 2.**
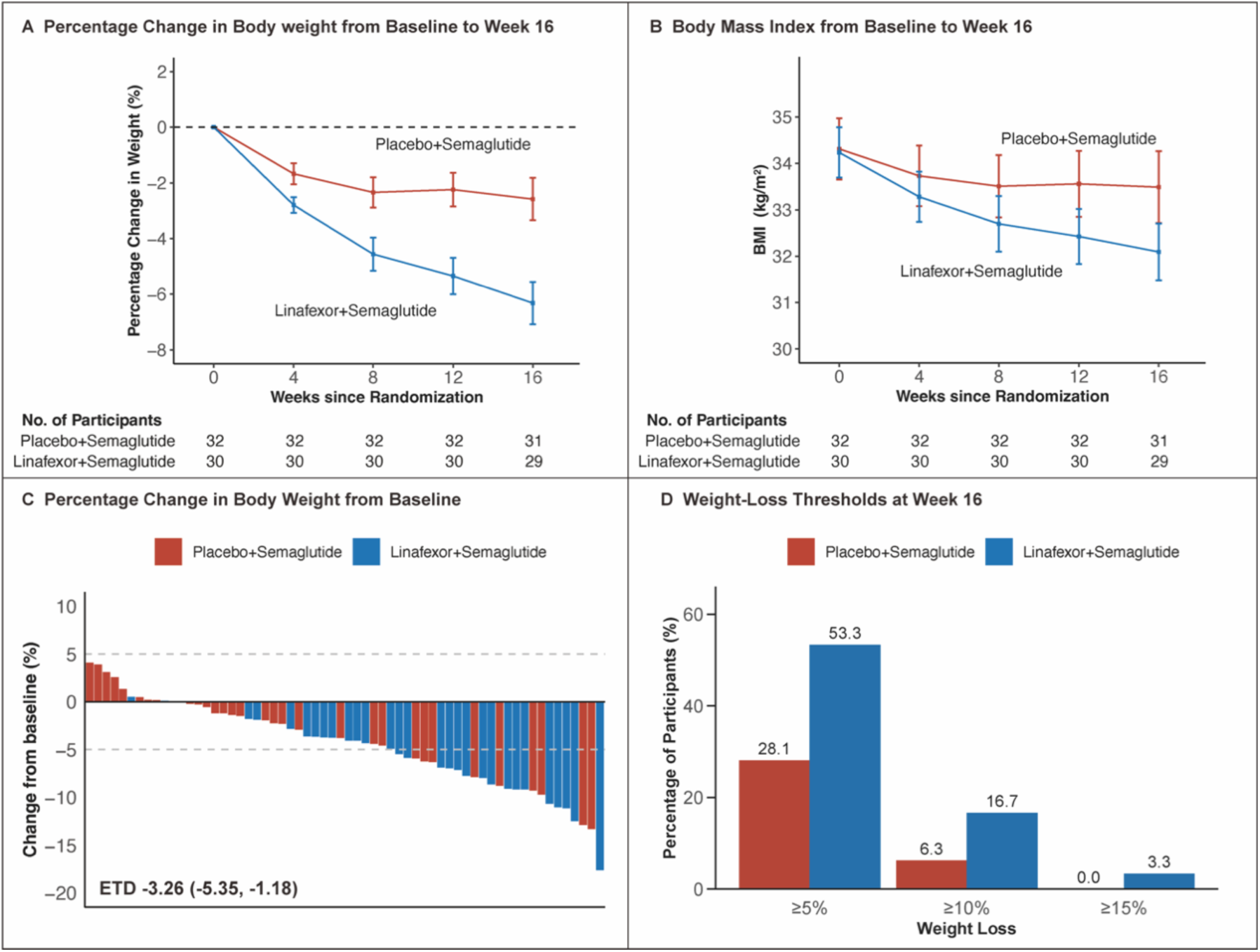
Effect of Linafexor plus Semaglutide, as Compared with Placebo plus Semaglutide, on Body Weight (Trial Product Estimand) Panel A shows the mean percentage change in body weight from baseline over the 16-week treatment period (the time from the first dose of the investigational product to the time of the last dose). Error bars indicate 95% confidence intervals. Panel B shows the mean change in body mass index (BMI) over time. Error bars indicate standard errors. In panel A and B, the numbers of participants with body weight measurements at various time points are shown. Panel C shows waterfall plot of the percentage change in body weight from baseline to week 16 for individual participants. Red bars indicate participants receiving placebo plus semaglutide, and blue bars indicate those receiving linafexor plus semaglutide. The horizontal dashed line at -5% represents the threshold for clinically meaningful weight loss. The Estimated Treatment Difference (ETD) was calculated using a mixed-effects model, incorporating treatment group, visit time, and their interaction as fixed effects. Panel D shows the observed percentages of participants who had body-weight reductions of at least 5%, at least 10%, and at least 15% from baseline to week 16.

**Table 2.**
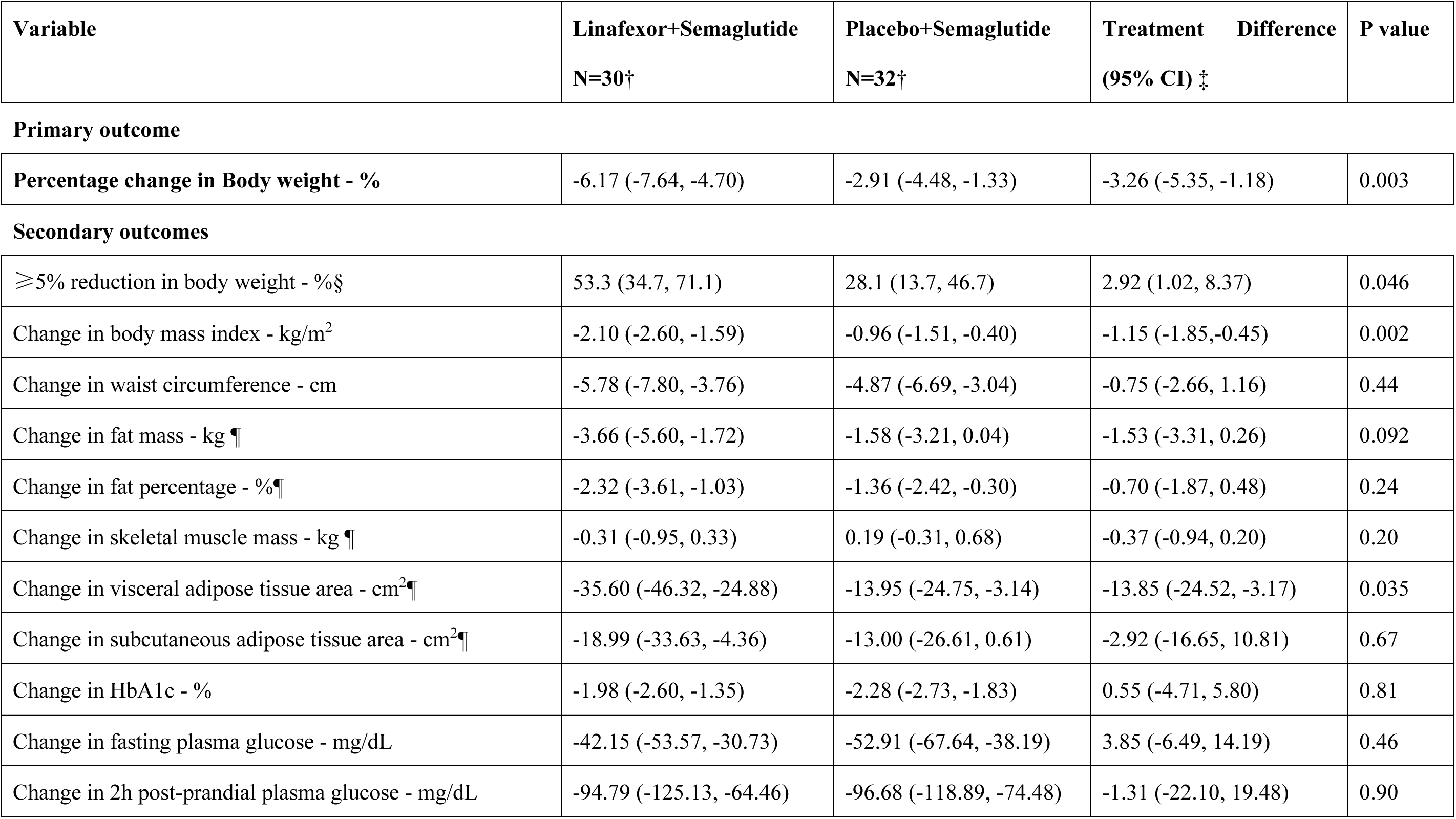
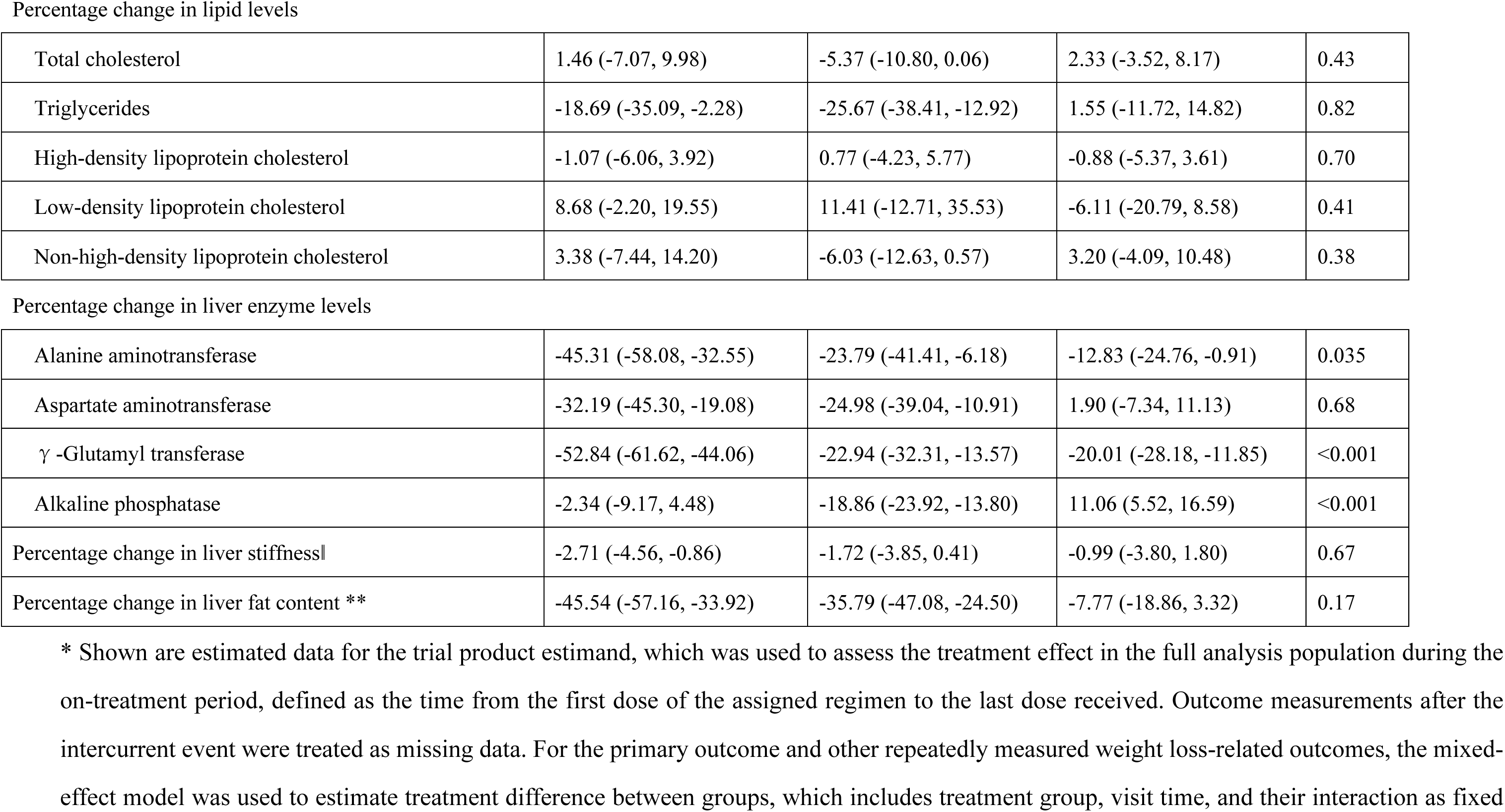
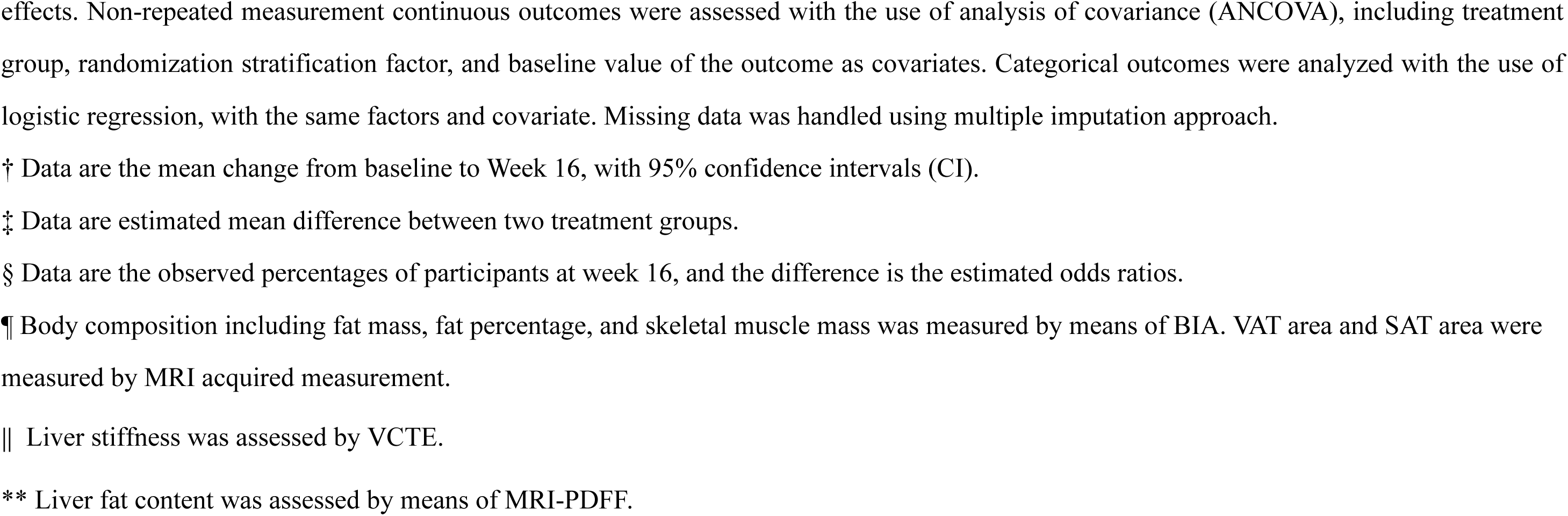
Primary and Key Secondary Outcomes at Week 16 (Trial Product Estimand).*

### Change in body composition

The additional weight lost with combination therapy was preferentially visceral fat. Although fat mass declined more in the linafexor plus semaglutide group than in the placebo plus semaglutide group (−3.66 kg vs. −1.58 kg), this difference was not statistically significant (**Table 2**). The contrast was clearer for visceral adipose tissue (VAT) area measured by MRI, which fell by −35.60 cm² (95% CI, −46.32 to −24.88) with combination therapy versus −13.95 cm² (95% CI, −24.75 to −3.14) with monotherapy—a treatment difference of −13.85 cm² (95% CI, −24.52 to −3.17; P=0.035) that strengthened to −16.8 cm² (95% CI, −27.3 to −6.3; P=0.002) after multivariable adjustment for covariates (**Table 2, Extended Data Table 3, and Fig. 3**). This greater fat loss was accompanied by minimal loss of skeletal muscle mass in the combination group (−0.31 kg), so that fat accounted for the large majority of the weight reduction.

**Fig 3.**
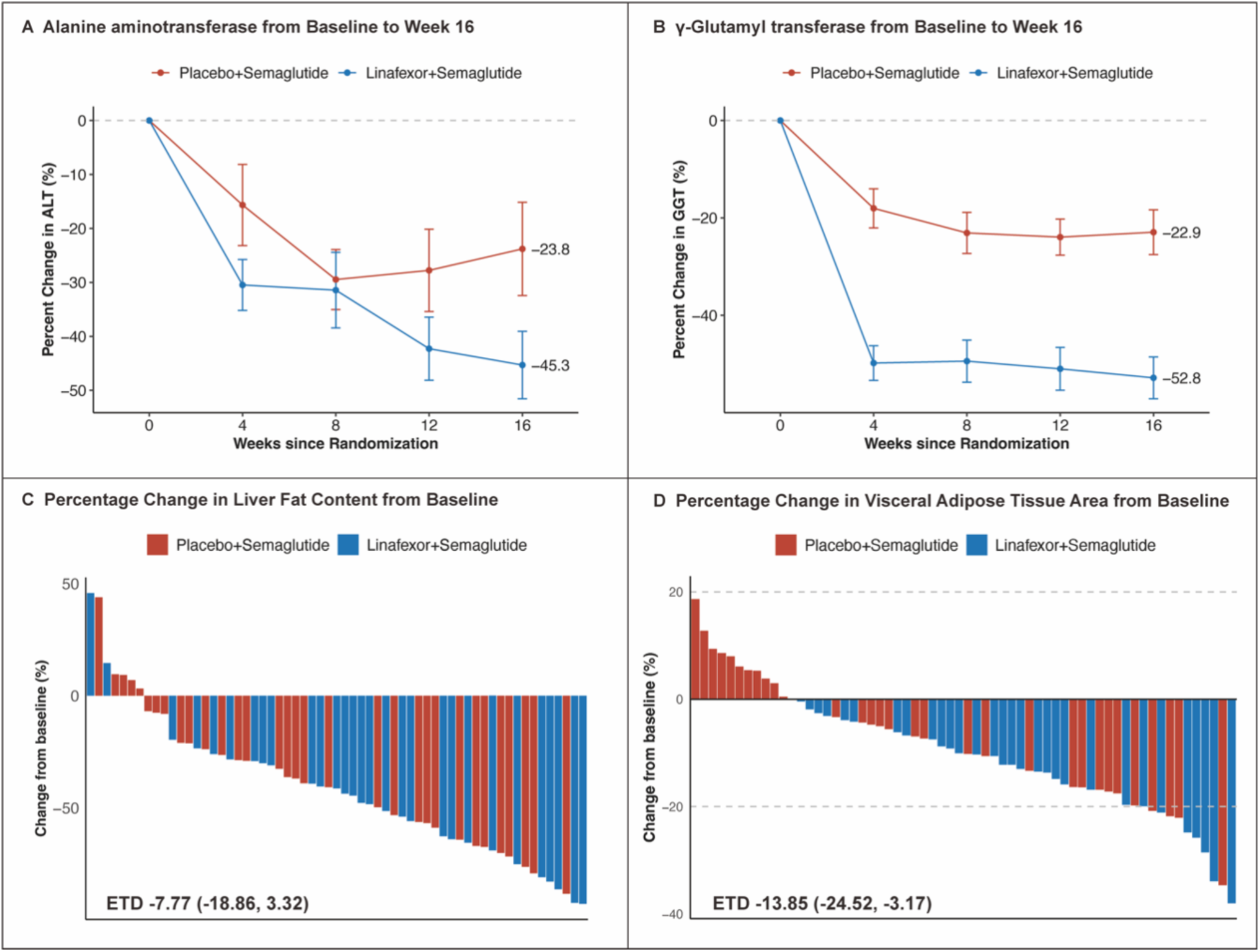
Changes in Liver enzymes, Liver fat content and Visceral adipose tissue area from baseline to Week 16. Panels A and B show mean percentage change in alanine aminotransferase (ALT) (A) and γ-Glutamyl transferase (GGT) (B) from baseline over the 16-week treatment period. Error bars indicate standard errors. Panels C and D show waterfall plots of the percentage change in liver fat content (C), and visceral adipose tissue area (D) from baseline to week 16 for individual participants. Red bars indicate participants receiving placebo plus semaglutide, and blue bars indicate those receiving linafexor plus semaglutide. The Estimated Treatment Difference (ETD) was calculated using analysis of covariance, incorporating treatment group, randomization stratification factor, and the baseline value of the outcome as covariates.

### Change in cardiometabolic risk factors

Improvements in glycemia and lipids were substantial but similar in the two groups, indicating that these benefits were driven largely by semaglutide rather than by linafexor. HbA1c fell by 1.98% (95% CI, −2.60 to −1.35) with combination therapy and by 2.28% (95% CI, −2.73 to −1.83) with monotherapy, with no significant between-group difference, and fasting and 2-hour postprandial plasma glucose improved comparably in both groups (**Table 2**). Triglyceride levels likewise decreased in both groups—by 18.69% (95% CI, −35.09 to −2.28) with combination therapy and 25.67% (95% CI, −38.41 to −12.92) with monotherapy—again without a significant difference between groups (**Table 2**).

### Change in hepatic parameters

In contrast to the glycemic and lipid measures, hepatic outcomes favored combination therapy. Liver fat content by MRI-PDFF declined by −45.54% (95% CI, −57.16 to −33.92) with combination therapy versus −35.79% (95% CI, −47.08 to −24.50) with monotherapy; although the unadjusted treatment difference was not significant (−7.77%; 95% CI, −18.86 to 3.32), multivariable adjustment revealed an additional 11.5% relative reduction in liver fat attributable to linafexor (95% CI, −21.5 to −1.5; P=0.025) (**Table 2 and Extended Data Table 3**). Liver stiffness by vibration-controlled transient elastography (VCTE) decreased significantly from baseline in the combination group (−2.71%; 95% CI, −4.56 to −0.86) but not in the monotherapy group (−1.72%; 95% CI, −3.85 to 0.41), though the between-group difference was not significant (−0.99%; 95% CI, −3.8 to 1.8) (**Table 2**). The clearest hepatic signal was in liver enzymes: alanine aminotransferase (ALT) fell by 45.31% (95% CI, −58.08 to −32.55) with combination therapy versus 23.79% (95% CI, −41.41 to −6.18) with monotherapy (between-group difference, −12.83%; 95% CI, −24.76 to −0.91), and γ-glutamyl transferase (GGT) fell by 52.84% (95% CI, −61.62 to −44.06) versus 22.94% (95% CI, −32.31 to −13.57), a between-group difference of −20.01% (95% CI, −28.18 to −11.85) (**Table 2 and Fig. 3**).

### Changes in bile acid profiles

To probe the mechanism underlying these hepatic benefits, we profiled serum bile acids (BAs) before and after the 16-week intervention. Fasting total BA levels diverged significantly between groups (**Fig. 4A**), falling in the linafexor plus semaglutide group (**Fig. 4B**) but rising in the placebo plus semaglutide group (**Fig. 4C**); detailed fasting and postprandial changes are summarized in Extended Data Tables 4 and 5. The two regimens reshaped the BA pool differently. In the linafexor plus semaglutide group, 13 BAs were differentially abundant in the fasting state (4 increased, 9 decreased) and 15 in the postprandial state (2 increased, 13 decreased), whereas in the placebo plus semaglutide group 13 BAs changed in the fasting state (5 increased, 8 decreased) but only 5 in the postprandial state (all increased). Linafexor produced a consistent pattern across both states—coordinated decreases in 3β-cholic acid (βCA), glycocholic acid (GCA), 3β-deoxycholic acid (βDCA), glycohyocholic acid (GHCA), glycochenodeoxycholic acid (GCDCA), and taurocholic acid (TCA), with increases in nordeoxycholic acid (NorDCA) and taurolithocholic acid (TLCA)— whereas monotherapy reduced BAs only in the fasting state (βDCA, GCDCA, and tauroursodeoxycholic acid [TUDCA]). These BA changes tracked with clinical response: in generalized estimating equation (GEE) analysis, reductions in GCA, βDCA, GHCA, GCDCA, and TCA were positively associated with the linafexor-induced reductions in body weight and liver enzymes, while NorDCA and TLCA showed the opposite direction of effect (**Fig. 4D and Fig. 4E**). No such BA-clinical correlations were evident with monotherapy (**Fig. 4F and Fig. 4G**).

**Fig 4.**
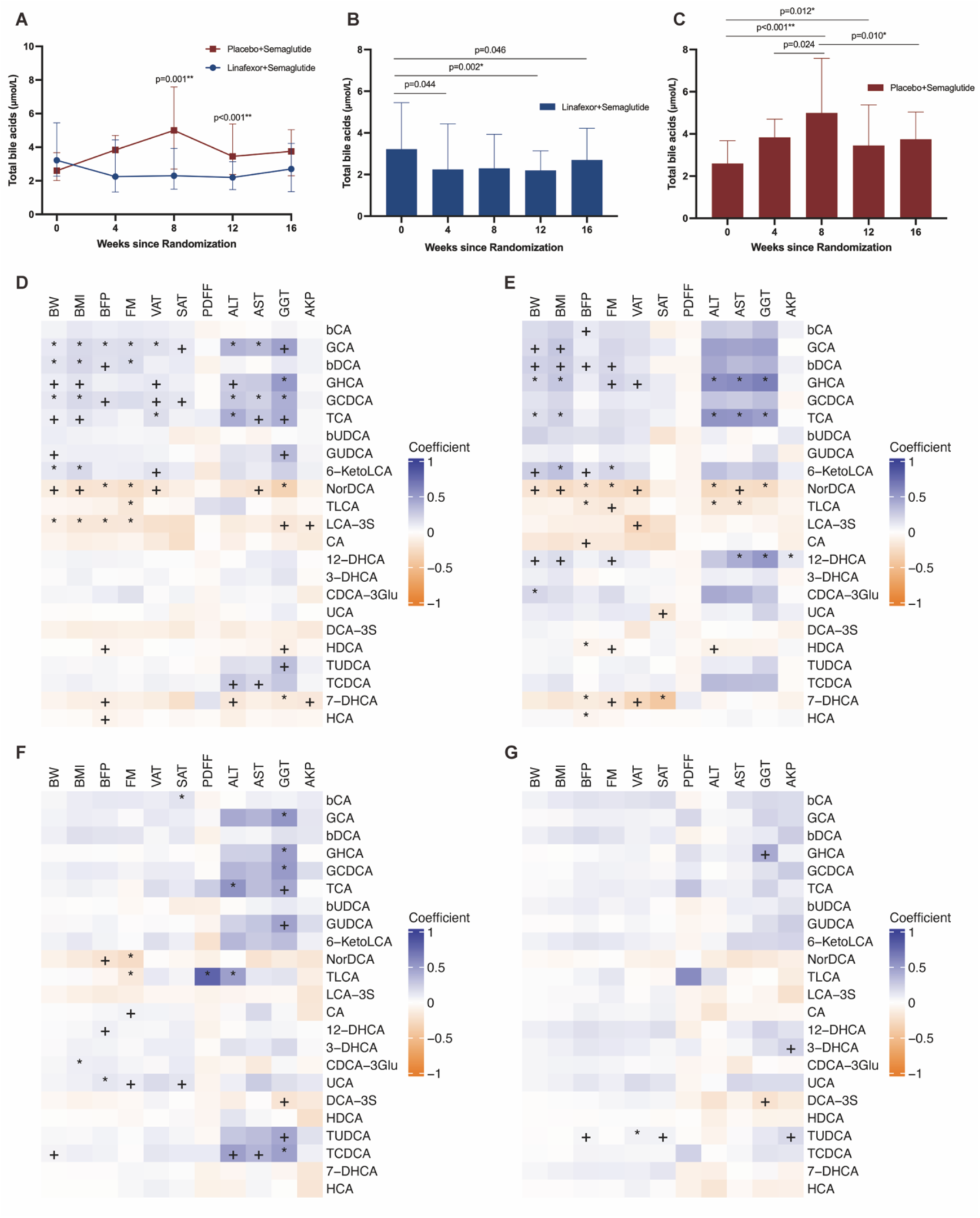
Changes in serum bile acid in response to treatment and correlations with clinical outcomes. Panel A shows the dynamic changes in fasting total bile acids (BAs) over the 16-week intervention between the linafexor plus semaglutide and placebo plus semaglutide groups. Data are presented as median and interquartile range (IQR). P-values indicate inter-group differences assessed by the Wilcoxon rank-sum test. Panels B and C show intra-group changes in fasting total BAs over the 16-week intervention for the linafexor plus semaglutide group (B) and the placebo plus semaglutide group (C). Data are presented as median and IQR. P-values indicate intra-group differences over time assessed by the Wilcoxon signed-rank test. In panels A-C, * False Discovery Rate (FDR)-adjusted P < 0.05, ** FDR-adjusted P < 0.01 (FDR correction via the Benjamini-Hochberg method). Panels D-G show heatmaps illustrating the correlations between specific BAs and clinical outcomes in the linafexor plus semaglutide group under fasting (D) and postprandial (E) states, and in the placebo plus semaglutide group under fasting (F) and postprandial (G) states. Correlations were evaluated using multivariate generalized estimating equations (GEE). The color gradient represents the correlation coefficient: blue indicates positive correlations, orange indicates negative correlations, and darker colors reflect stronger correlations. In panels D-G, + FDR-adjusted P < 0.05, * FDR-adjusted P < 0.01 (FDR correction via the Benjamini-Hochberg method). βCA, 3β-Cholic Acid; GCA, Glycocholic Acid; βDCA, 3β-Deoxycholic Acid; GHCA, Glycohyocholic Acid; GCDCA, Glycochenodeoxycholic Acid; TCA, Taurocholic Acid; βUDCA, Isoursodeoxycholic Acid; GUDCA, Glycoursodeoxycholic Acid; 6-KetoLCA, 6-Ketolithocholic Acid; NorDCA, Nordeoxycholic Acid; TLCA, Taurolithocholic Acid; LCA-3S, Lithocholic Acid 3-Sulfate; CA, Cholic Acid; 12-DHCA, 12-Dehydrocholic Acid; 3-DHCA, 3-Oxocholic Acid; CDCA-3Glu, Chenodeoxycholic Acid-3-β-D-Glucuronide; UCA, Ursocholic Acid; DCA-3S, 3-Sulfodeoxycholic Acid; HDCA, Hyodeoxycholic Acid; TUDCA, Tauroursodeoxycholic Acid; TCDCA, Taurochenodeoxycholic Acid; 7-DHCA, 7-Dehydrocholic Acid; HCA, Hyocholic Acid;

### Pharmacokinetic profile

To confirm that linafexor retained its intended pulsatile exposure in this population, we performed pharmacokinetic (PK) sampling in a subset of participants at week 16. In the linafexor plus semaglutide group (n=9), oral linafexor 4 mg was rapidly absorbed, reaching peak plasma concentrations at approximately 0.5 hours post-dose, and was then eliminated extremely rapidly, with concentrations declining sharply toward the lower limit of quantification (LLOQ) shortly thereafter (**Extended Data Fig. 3**). All PK values in the placebo plus semaglutide group (n=10) were below the LLOQ. This brief, transient exposure confirms pulsatile rather than sustained FXR engagement and is consistent with the predominantly postprandial bile-acid changes described above.

### Safety and tolerability

The combination was well tolerated, with a safety profile comparable to that of low-dose semaglutide alone. Adverse events were reported in 70.0% of the linafexor plus semaglutide group and 56.2% of the placebo plus semaglutide group; most were mild, and none led to treatment discontinuation (**Table 3**). No participant discontinued semaglutide or required a dose reduction from 0.5 mg to 0.25 mg weekly because of adverse events. Gastrointestinal events—the principal dose-limiting toxicity of GLP-1 receptor agonists—were infrequent and similar between groups (23.3% with combination therapy vs. 18.8% with monotherapy), with comparable time to first onset (**Table 3 and Extended Data Fig. 4**); two episodes of vomiting occurred in the monotherapy group (2/32) and none in the combination group (0/30). Pruritus, a class effect of FXR agonists, was reported in only two participants receiving combination therapy and one receiving monotherapy, all mild and self-limited, and no participant developed elevated alkaline phosphatase or bilirubin—distinguishing linafexor from earlier FXR agonists. Hypoglycemic episodes were infrequent and mild in both groups, with no severe hypoglycemia. Four serious adverse events occurred—three in the combination group (duodenitis requiring hospitalization, and colorectal and gastric polyps requiring endoscopic removal during hospitalization) and one in the control group (induced abortion)—but all occurred after the 16-week treatment period and none were considered treatment-related by the investigators. Laboratory monitoring revealed no signal of renal dysfunction, electrolyte disturbance, or hepatotoxicity.

**Table 3.**
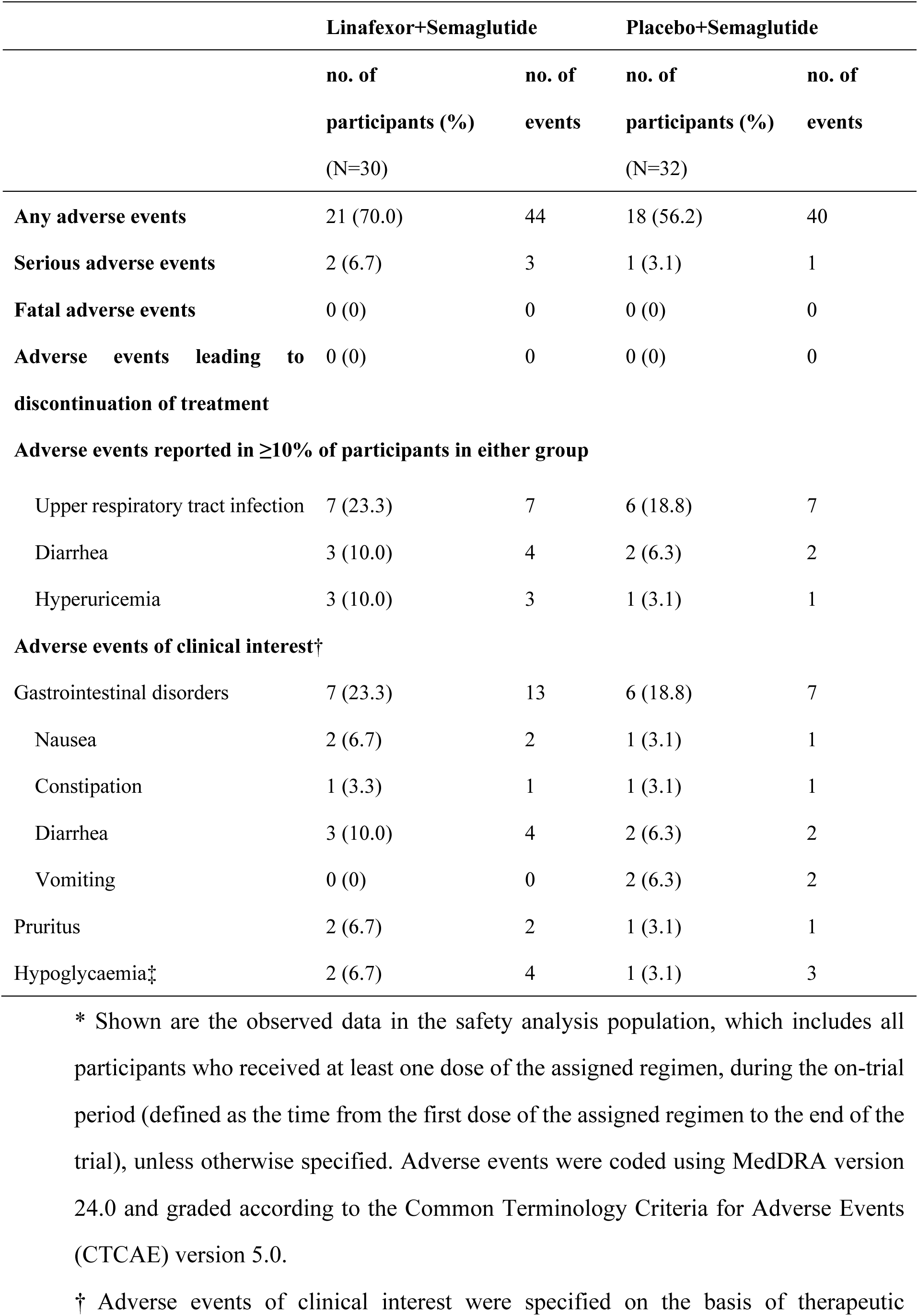
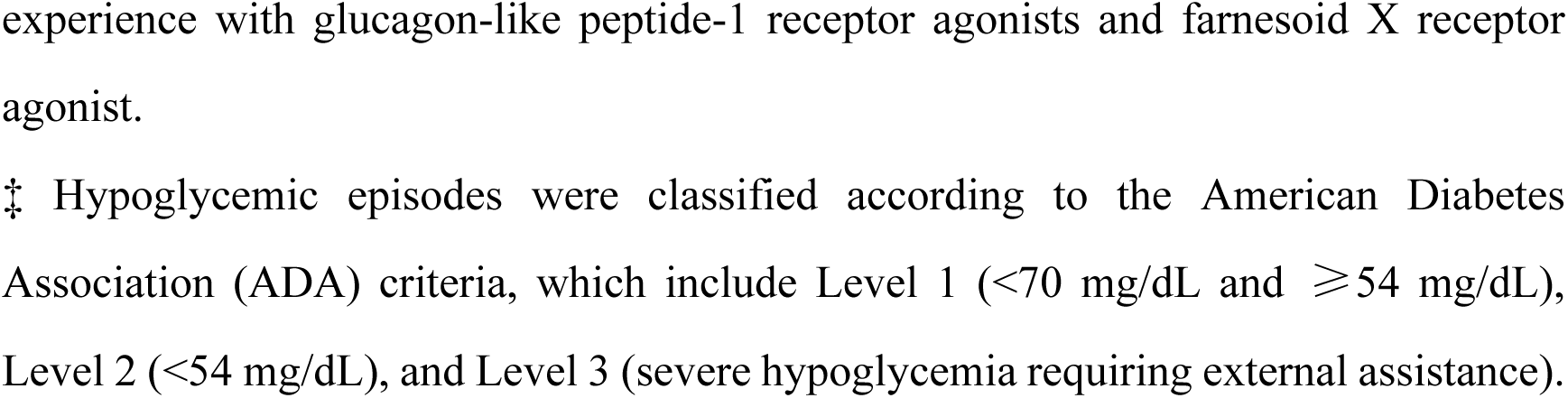
Adverse Events.*

## Discussion

In this multicenter, randomized, double-blind, placebo-controlled phase 2 trial, we provide what is, to our knowledge, among the first clinical evidence that adding linafexor, a pulsatile FXR agonist, to low-dose semaglutide (0.5 mg weekly) produces significantly greater weight loss than low-dose semaglutide alone over 16 weeks in adults with obesity, MASH, and type 2 diabetes—a population in whom the weight-loss response to GLP-1 receptor agonists is characteristically blunted. Beyond weight, the combination conferred greater visceral fat reduction, larger improvements in liver fat and liver enzymes, and a distinct, clinically correlated shift in the bile acid pool, all while maintaining tolerability indistinguishable from low-dose semaglutide alone. Together, these findings establish proof of concept that pairing complementary mechanisms can enhance efficacy in a difficult-to-treat population without incurring the dose-dependent toxicity that limits GLP-1 monotherapy.

The principal advance demonstrated here is in tolerability. High-dose GLP-1 receptor agonists have transformed obesity treatment, but their use is constrained by gastrointestinal intolerance affecting up to 63–74% of patients^4,13^, frequent treatment discontinuation, and high cost, as well as by loss of lean mass^29^. By combining linafexor with just 20% of the standard semaglutide obesity dose, we achieved clinically meaningful weight loss while largely circumventing these liabilities: gastrointestinal adverse events occurred in only 23.3% of the combination group, no participant discontinued treatment because of adverse events, and none required a semaglutide dose reduction. A 6.17% mean weight loss—of a magnitude ordinarily associated with substantially higher semaglutide doses^10,13^—indicates that linafexor permits the semaglutide dose, and its attendant toxicity, to be markedly reduced without sacrificing efficacy.

The combination also produced clear hepatic benefit. Liver fat content fell by 45.54% with combination therapy—exceeding what is typically achieved with pharmacotherapy over comparable periods—and although the between-group difference reached significance only after multivariable adjustment, it was accompanied by disproportionate reductions in the liver enzymes ALT and GGT that are unlikely to be explained by weight loss alone. These enzyme changes point to a direct hepatoprotective effect of FXR activation. Because MASH arises from intertwined metabolic, inflammatory, and fibrotic processes, simultaneously engaging the GLP-1 and FXR pathways offers complementary anti-inflammatory and anti-fibrotic actions^30–35^, addressing obesity and MASH—conditions that frequently co-occur and share metabolic roots—within a single regimen. These results provide proof of concept for dual-pathway intervention in metabolic liver disease.

Body composition outcomes further favored the combination, with a preferential loss of visceral adipose tissue. Because visceral adiposity is more strongly linked to insulin resistance and cardiovascular risk than subcutaneous fat, its greater reduction in the combination group may carry cardiometabolic benefit beyond the weight change itself. Notably, the combination group lost fat and skeletal muscle in an approximately 12:1 ratio (−3.66 kg fat vs. −0.31 kg skeletal muscle mass), suggesting that FXR agonism may favorably shift nutrient partitioning during weight loss and help preserve lean mass—an important consideration given the muscle loss associated with high-dose GLP-1 therapy.

These complementary clinical effects are consistent with the distinct mechanisms of the two agents. Semaglutide acts principally through appetite suppression and delayed gastric emptying^36^, whereas pulsatile FXR activation by linafexor engages distinct metabolic pathways—increased energy expenditure, improved insulin sensitivity, and favorable nutrient partitioning—without the receptor desensitization and toxicity that accompany continuous FXR stimulation. This mechanistic complementarity provides a plausible basis for the dose-sparing synergy observed here and may explain how the combination overcame the attenuated GLP-1 response typical of obese diabetic patients, potentially through improved insulin signaling, enhanced mitochondrial function, and dampened inflammation^37–39^.

Serum bile acid profiling offered a molecular window onto these effects. In non-alcoholic fatty liver disease (NAFLD), circulating bile acids are paradoxically elevated despite suppressed hepatic FXR signaling^40^. Consistent with this, semaglutide monotherapy further increased total bile acids in our trial, suggesting that its hepatic benefits arise from broad metabolic effects rather than from correcting bile acid dysregulation^31^. Linafexor produced the opposite effect: by activating FXR and suppressing bile acid synthesis, it markedly lowered total bile acids, and the combination group showed the broadest downregulation, most pronounced in the postprandial state. This postprandial predominance likely reflects linafexor’s short half-life, whereby pre-meal dosing blunts the postprandial surge in circulating bile acids. Critically, these molecular changes tracked with clinical response: reductions in GCA, βDCA, GCDCA, GHCA, and TCA correlated with improvements in body weight and liver enzymes, whereas NorDCA and TLCA moved in the opposite direction. This directionality is biologically coherent—GCA, GCDCA, and TCA are elevated in MASH, and GCDCA has been proposed as a biomarker of liver injury^41,42^; NorDCA is reduced in chronic liver disease and in murine NAFLD livers^43,44^; and TLCA is an endogenous agonist of Takeda G protein-coupled receptor 5 (TGR5), whose activation promotes energy expenditure, suppresses inflammation, and modulates appetite^45^, with TGR5 agonists ameliorating diet-induced steatosis and insulin resistance in animal models^46^. Collectively, these data indicate that adding linafexor reverses the dysregulated bile acid profile that characterizes MASH.

A further observation reinforces the rationale for this combination in hard-to-treat patients. Whereas semaglutide’s weight-loss efficacy is generally greatest in non-diabetic individuals and in those with lower baseline HbA1c^4,13,47^, the linafexor– semaglutide combination produced its most pronounced weight loss in participants with HbA1c >8.5% (**Extended Data Fig. 2**). Although the interaction did not reach statistical significance, the direction of effect is the inverse of what is observed with semaglutide alone and suggests that the combination may specifically benefit the patients least responsive to GLP-1 monotherapy—a hypothesis that warrants confirmation in adequately powered trials.

From a practical standpoint, these findings have implications for treatment access. Achieving meaningful weight loss at one-fifth of the standard semaglutide dose could lower drug costs by roughly 80% while improving treatment persistence, and the combination’s superior tolerability and apparent preservation of skeletal muscle broaden its applicability—particularly for patients prone to the adverse effects that drive discontinuation of high-dose GLP-1 therapy^48^ and for those at risk of sarcopenia^12,49^. Such a regimen could expand access to effective obesity treatment.

This trial has several strengths, including its randomized, double-blind design and high retention (>96%), which support the validity of the findings, and the consistency of results across multiple sensitivity analyses. The paired pre-and post-treatment bile acid profiling and the on-treatment pharmacokinetic sampling add a degree of mechanistic depth uncommon in early-phase efficacy trials.

Several limitations temper interpretation. First, by design the comparator was low-dose (0.5 mg) semaglutide rather than a standard obesity dose, so the trial cannot establish how the combination compares with full-dose semaglutide—the comparison most relevant to clinical practice. Second, MASH was defined by MRI-PDFF ≥10% rather than by liver biopsy; this is appropriate for a phase 2 trial but falls short of histologic confirmation. Third, the 16-week duration was relatively short: weight loss had not plateaued, so the full between-group difference may not yet have emerged, and the interval was likely too brief to detect histologic change or a difference in liver stiffness. Longer, adequately powered trials—ideally including a standard-dose semaglutide comparator and histologic endpoints—are needed to confirm and extend these findings.

In conclusion, adding the pulsatile FXR agonist linafexor to low-dose semaglutide more than doubled weight loss while improving body composition and hepatic measures and preserving the tolerability of low-dose therapy, in a population in which GLP-1–induced weight loss is characteristically blunted. By coupling complementary mechanisms to gain efficacy without dose-dependent toxicity, this dose-sparing strategy addresses central limitations of current obesity pharmacotherapy and offers a rational template for combination treatment of obesity and its metabolic complications.

## Methods

### Study design

This multicenter, randomized, double-blind, placebo-controlled phase 2 trial was designed to evaluate the efficacy and safety of linafexor in combination with low-dose semaglutide for weight management in obese adults with metabolic dysfunction-associated steatohepatitis (MASH) and type 2 diabetes. This trial was conducted at four sites in China, which included a 2-week screening period, a 16-week treatment period, and a 4-week off-treatment follow-up period (**Extended Data Fig. 1**). This trial has been registered at ClinicalTrials.gov (NCT06492330), was approved by the ethics committee at Ruijin Hospital and at each participating site, and conducted in accordance with the Declaration of Helsinki and Good Clinical Practice guidelines. All participants provided written informed consent before enrollment. Participants and the public were not involved in the design, conduct and reporting of the trial

### Participants and eligibility criteria

Eligible participants were adults aged 18 to 65 years with obesity, defined as a body mass index (BMI) between 30 and 45 kg/m^2^. Participants were required to have both type 2 diabetes (HbA1c level of 7.0% to 10.5% and fasting plasma glucose of 126 to 240 mg/dl at screening) and MASH. MASH was confirmed by either a liver biopsy, or by magnetic resonance imaging proton density fat fraction (MRI-PDFF) ≥10% within 3 months prior to randomization. Blood glucose had been controlled solely through lifestyle interventions (diet and exercise) for at least 3 months prior to screening. Key exclusion criteria included weight change greater than 5% within 3 months or by more than 10% within 6 months before randomization; use any of weight-loss medications, or use of glucagon-like peptide-1 (GLP-1) receptor agonists and any other hypoglycemic medications within 3 months prior to randomization; diseases that interfere with the drug absorption, distribution, metabolism or excretion including inflammatory bowel disease, or previous gastric bypass surgery; decompensated liver disease, hepatitis B or C infection; known drug abuse or excessive alcohol consumption. Details of the inclusion and exclusion criteria are provided in the study protocol in the Supplementary Information.

### Randomization, blinding and treatment allocation

Participants were randomly assigned in a 1:1 ratio to receive either linafexor plus semaglutide or matching placebo plus semaglutide. Randomization was performed centrally using an interactive web response system with a computer-generated sequence. Randomization was stratified by baseline BMI (<35 or ≥35 kg/m²). Trial-group assignments were concealed from participants, investigators, site staff, and sponsor personnel involved in trial conduct and data analysis until database lock. Linafexor and matching placebo tablets were identical in appearance, taste, and packaging. Semaglutide was administered using prefilled pen injectors with the dose clearly labeled; both groups received identical training on injection technique.

### Interventions

After a 2-week screening period, eligible participants were randomly assigned to receive either oral linafexor (4 mg once daily; Cascade Pharmaceuticals, Shanghai, China) plus subcutaneous semaglutide (0.5 mg once weekly; Novo Nordisk, Bagsværd, Denmark) or matching oral placebo plus subcutaneous semaglutide (0.5 mg once weekly) for 16 weeks, followed by a 4-week off-treatment follow-up period. All participants received standardized lifestyle counseling (diet and exercise) at baseline and every 4 weeks throughout the trial (details in the study protocol in the Supplementary Information). Lifestyle recommendations focused on healthy, balanced meals aiming to reduce total daily caloric intake by at least 400–500 kcal, combined with at least 20 minutes daily or 150 minutes per week of moderate-intensity aerobic exercise. Dose adjustments for semaglutide were permitted only under predefined conditions. If participants experienced recurrent symptoms of hypoglycemia with confirmed fasting plasma glucose ≤ 70 mg/dL, or intolerable gastrointestinal adverse events deemed related to the investigational product by the investigator, the semaglutide dose was reduced to 0.25 mg once weekly. If fasting plasma glucose was ≥ 240 mg/dL and confirmed upon re-testing within 3 days, the semaglutide dose was increased to 1.0 mg once weekly. Each participant was permitted only one dose reduction due to hypoglycemia or gastrointestinal intolerance, and one dose increase due to hyperglycemia. Participants were discontinued from the study if glycemic control became unacceptable again or if the same intolerable gastrointestinal symptoms recurred. Participants attended site visits at weeks 4, 8, 12, and 16 (end of treatment), during which protocol-specified assessments were performed, and adverse events were recorded. Safety evaluations were also performed at week 20 (end of follow-up). After providing additional written informed consent, a subset of participants underwent pharmacokinetic blood sampling before and after dosing at the Week 16 visit.

## Outcomes

### Primary outcomes

The primary outcome was the percentage change in body weight from baseline to week 16.

### Weight measurement

Body weight was measured in the morning after participants had fasted, emptied their bladder, and removed footwear and socks, wearing a single light layer of clothing. Participants stood still on a calibrated scale placed on a stable, level, hard surface, and weight was recorded to the nearest 0.1 kg once the reading stabilized. Two investigators cross-checked each value, and the same scale was used throughout the study.

### Secondary outcome

Key secondary outcomes included the proportion of participants achieving weight loss of at least 5% at week 16; changes in body weight and BMI; changes in waist circumference; changes in body composition (fat mass, skeletal muscle mass, percentage body fat) measured by bioelectrical impedance analysis (BIA); changes in visceral adipose tissue (VAT) and subcutaneous adipose tissue (SAT) area were measured by MRI; changes in liver stiffness was measured by vibration-controlled transient elastography (VCTE); changes in glycemic control (including HbA1c, fasting and postprandial 2 hour plasma glucose); changes in liver enzyme levels (alanine aminotransferase [ALT], aspartate aminotransferase [AST], γ-glutamyl transferase [GGT], alkaline phosphatase [ALP]); changes in liver fat content measured by MRI-PDFF; changes in lipid profiles; all evaluated as changes from baseline to week 16.

### Waist and hip circumference measurement

Waist and hip circumference measurements were performed by trained investigators under fasting conditions and recorded to one decimal place (cm). Participants stood upright without outer garments or belts, with feet shoulder-width apart and arms relaxed at their sides. A nonelastic, flexible measuring tape was placed horizontally (parallel to the floor), snug but without compressing the skin, with consistent tension. Waist circumference was measured midway between the lower costal margins and the iliac crests at end-expiration in a standing position. Hip circumference was measured at the level of the greatest posterior protuberance of the buttocks (around the greater trochanter) in a standing position.

### Body composition measurement

Body composition was measured by bioelectrical impedance analysis (BIA) to determine fat mass, body fat percentage, and skeletal muscle mass, using the InBody 770 or 720 device (InBody Co., Seoul, Korea), which applies multiple electrical frequencies to improve body composition estimation^50,51^. Participants were measured under fasting conditions (no food or fluids for ≥8 hours), with an empty bladder, and having avoided vigorous exercise for 24 hours prior. Before the procedure, participants sat or stood quietly for 5 minutes. Shoes, socks, jewelry, and watches were removed, pockets emptied, and participants wore light clothing. Hands and soles were ensured clean and dry, with wipes used if necessary for better contact. The device was placed on a stable, level, hard surface. Participants stood barefoot on the foot electrodes, aligned their heels to the device markers, and grasped the hand electrodes per device instructions with elbows extended and arms slightly abducted (∼45°) from the trunk, remaining still and silent throughout. All relevant parameters were displayed on the device screen, and a trained investigator supervised and standardized the procedure and recorded the results.

### Liver fat content measurement

Liver fat content was measured using magnetic resonance imaging proton density fat fraction (MRI-PDFF), which quantitatively assesses liver fat content and reports results as a percentage. Scanning was performed on the same MRI scanner at each site for all visits. After image acquisition, the system automatically reconstructed and output PDFF parametric maps, which were used as the source for quantitative results. Participants fasted for 4–6 hours before the examination, avoided strenuous exercise and alcohol for approximately 24 hours, and completed standard MRI safety screening prior to the procedure. During the scan, participants were positioned supine, and imaging covered the entire liver. Image acquisition was performed under breath hold whenever feasible. To ensure consistency, the same technologist at each site conducted the examinations and quantitative readings whenever possible, avoiding major vessels, bile ducts, obvious artifacts, and focal lesions.

### Visceral and subcutaneous adipose tissue measurements

Visceral adipose tissue (VAT) and subcutaneous adipose tissue (SAT) were manually delineated using ITK-SNAP segmentation software (version 3.6) on axial PDFF images. The assessment of abdominal fat was conducted at the L1-L2 level, excluding intestinal loops^52^. VAT was characterized as intraabdominal fat, parietal peritoneum or transversalis fascia, while excluding the vertebral column and paraspinal muscles^53^. SAT was defined as the adipose tissue located superficial to the abdominal and dorsal musculature^54^.

Quantitative analysis was conducted using Python (version 3.6, Python Software Foundation). The VAT and SAT areas (cm^2^) were calculated as the sum of pixels within each segmented region multiplied by the pixel size. A threshold of 50% of the peak fat signal intensity was employed; pixels exceeding this threshold were classified as adipose tissue for quantifying SAT and VAT areas^55^.

### Safety outcomes

Safety outcomes included adverse events (AEs), serious adverse events (SAEs), discontinuations due to adverse events, hypoglycemic episodes (graded according to American Diabetes Association criteria), physical examinations, vital signs, 12-lead electrocardiogram (12-ECG), psychiatric symptoms, and changes in laboratory parameters including blood routine, urine routine and blood biochemistry.

### Bioanalytical methods for pharmacokinetic samples

Chromatographic separation was performed using ExionLC™ AD (AB Sciex, USA) on ACQUITY UPLC BEH C18 analytical column (2.1 × 50 mm, Waters, USA) at 40°C. The mobile phases were 1 mM ammonium formate with 0.02% formic acid in water (A) and acetonitrile (B). The flow rate was 0.6 mL/min, and the run time was 4 minutes. A Triple Quad QTRAP 6500+ mass spectrometer (AB Sciex, USA) coupled with an electrospray ionization ion source was used to monitor the mass transitions. The system was operated in positive mode, and multiple reaction monitoring mass transitions were m/z 571.1/527.1 and m/z 579.1/535.1 for Linafexor and its internal standard (IS), respectively. A 100-μL volume of human plasma and 20-μL volume of IS working solution were added into a 2-mL centrifuge tube. Then a 50-μL volume of modifier was added. Mix well and 500-μL volume of ethyl acetate was used to extract Linafexor. Thereafter, the supernatant liquid was transferred and dried with air at 45℃. Then a 400-μL volume of acetonitrile: water (50:50, v/v) was used to reconstitute the residue. At last, 5 μL of sample solution was injected onto the liquid chromatography with tandem mass spectrometry (LC-MS/MS) system. The standard curve is from 0.1 ng/mL to 80.0 ng/mL.

### Bile acid profiling of human serum

A targeted panel of 66 serum bile acids (BAs) was quantified before and after the intervention. The complete list of analyzed BAs can be found in the Supplementary Information. Bile acid analysis was provided and approved by Metabo-Profile Biotechnology (Shanghai) Co., Ltd. All bile acid (BA) standards were purchased from Steraloids Inc. (Newport, RI, USA), TRC Chemicals (Toronto, ON, Canada), or custom synthesized by MP laboratory. Six stable isotope-labeled standards were obtained from C/D/N Isotopes Inc. (Quebec, Canada) and Steraloids Inc. Ammonium acetate was purchased from Sigma-Aldrich (St. Louis, MO, USA). Methanol, isopropyl alcohol, acetonitrile, glacial acetic acid, and formic acid were obtained from Thermo-Fisher Scientific (FairLawn, NJ, USA). For sample preparation, 50 μL of serum was added to a 96-well plate and mixed with 400 µL of acetonitrile/methanol (8:2, v/v) containing six internal standards. After centrifugation at 650 rpm for 20 min at 10 °C, 250 μL of the supernatant was transferred to a clean plate and lyophilized.

The dried samples were reconstituted in 40 μL of acetonitrile/methanol (80:20, v/v) and centrifuged at 650 rpm for 20 min at 10 °C. Subsequently, 60 µL of deionized water was added to the plate, stored at -20 °C for 20 min, and centrifuged at 4000×g for 30 min at 4 °C. Quantitative analysis was performed using an ACQUITY UPLC-Xevo TQ-S system (Waters Corp., Milford, MA, USA) with an injection volume of 5 µL. Pooled quality control (QC) samples were injected after every 14 test samples to monitor analytical variations. Raw UPLC-MS/MS data were processed using MassLynx software (v4.1, Waters) for peak integration and quantitation. Statistical analyses were performed using the iMAP platform (v1.0, Metabo-Profile, Shanghai, China).

## Sample size

The sample size calculation was based on the primary outcome. Assuming a standard deviation of 2.5% for percentage weight change in both groups and two-sided alpha of 0.05, 25 participants per group would provide 80% power to detect a 2% difference between groups. Accounting for a 15% dropout rate, we planned to enroll 30 participants per group.

## Statistical analysis

The primary analysis population included all randomized participants (intention-to-treat principle). Treatment efficacy was assessed using two estimands: the trial product estimand and the treatment policy estimand. The trial product estimand, which is the primary estimand of this study, assessed the efficacy during the on-treatment period before intercurrent events (early discontinuation of the study treatment or initiation of rescue therapy). For trial product estimand, the primary outcome was analyzed using mixed-effect model including treatment group, visit, and treatment-by-visit interaction as fixed effects. The treatment policy estimand assessed the efficacy regardless the intercurrent events. For treatment policy estimand, the primary outcome was analyzed using analysis of covariance (ANCOVA). Missing data will be handled using multiple imputation (MI), performed separately in the two treatment groups.

Sensitivity analysis on Per-Protocol set and respecified subgroup analysis for age, sex, BMI, liver fat content, HbA1c, ALT at baseline were also performed for primary outcomes. Post-hoc multivariate adjusted analysis was conducted for liver fat content and VAT area as exploratory analysis.

Generalized estimating equation (GEE) analysis was performed to assess the associations between changes in bile acid species and clinical parameters in both treatment arms. P values for each regression coefficient were calculated and adjusted for false discovery rate (FDR) using the Benjamini-Hochberg method, with a q value <0.05 considered statistically significant.

All statistical analysis was performed using R (version 4.3). Unless otherwise specified, all statistical tests will be two-sided with a significance level of 0.05. No multiplicity adjustment will be made for secondary outcomes. Detailed statistical analysis plans (SAP) are provided in the Supplementary Information.

## Data availability statement

The study data can be requested from the corresponding author, Weiqing Wang, 6 months after its publication.

## Acknowledgments

We thank all the patients, investigators, and trial staff members who were involved in the conduct of the trial.

## Author contributions

W.W., H.X., G.N., G.Z. and G.W. designed the trial; W.Z. and M.Q. performed data analysis; H.X., Y.Z. and Y.C. interpreted the data; Y.Z., Y.C., S.H., L.J., Y.W., J.M., J.D., L.X., Y.M., P.W. and C.L. conducted the trial and collected data; H.L. and F.Y. processed MRI imaging; H.X., Y.Z., Y.C. and R.D. drafted the manuscript; Y.Z., Y.C., H.X., G.N. and W.W. critically revised the work. W.W., H.X., G.N. and Y.Z. had full data access. All authors approved the final version, assume responsibility for submission decisions, and vouch for data accuracy and protocol compliance.

