## Supplementary Material for "Linafexor, a Pulsatile FXR Agonist, Augments Low-Dose Semaglutide for Weight Loss in Adults with Obesity, MASH, and Type 2 Diabetes"

**Supplementary Appendix - Table of Contents**

**Extended Data Fig. 1 |** Trial Design…………………………………………….……..…......2

**Extended Data Fig. 2 |** Primary Outcome According to Prespecified Subgroup.........3

**Extended Data Fig. 3 |** Plasma Concentration-Time Profiles of Linafexor at Week 16………...................................................................................................................4

**Extended Data Fig. 4 |** Time to Onset of First Gastrointestinal Adverse Event...........5

**Extended Data Table 1 |** Primary and Key Secondary End Points at Week 16 (Treatment Policy Estimand)……………………………………………………….........6

**Extended Data Table 2 |** Primary and Key Secondary End Points at Week 16 (Per-Protocol Set) …...………………………………………………………………...........8

**Extended Data Table 3 |** Association Between Intervention and Changes in Liver fat content and Visceral Adipose Tissue Area……………….........................................10

**Extended Data Table 4 |** Differential bile acids before and after 16-week intervention in the Linafexor plus Semaglutide group………...............................................……..11

**Extended Data Table 5 |** Differential bile acids before and after 16-week intervention in the Placebo plus Semaglutide group………........................……………..…….13

**Extended Data Fig. 1 | Trial Design**

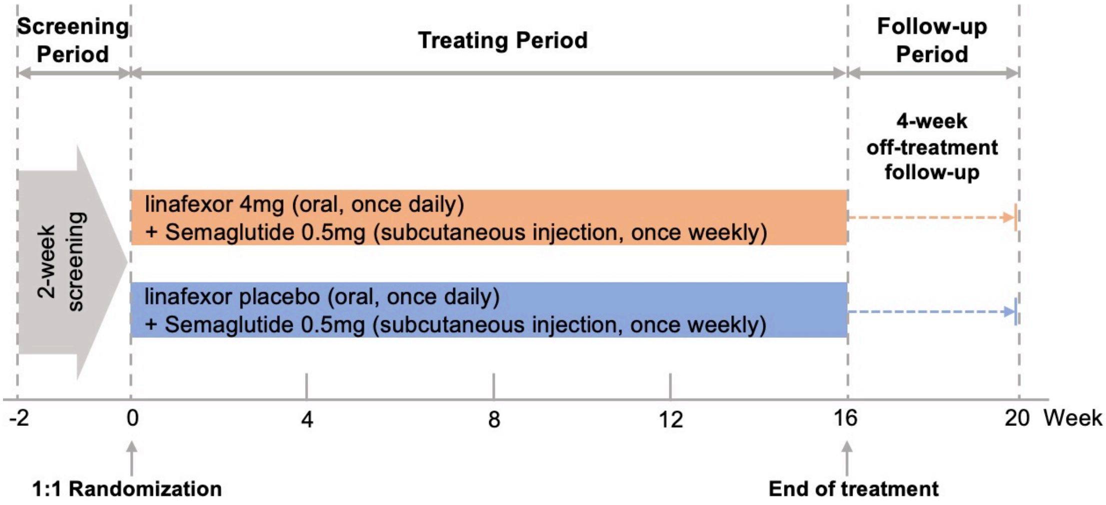


This multicenter, randomized, double-blind, placebo-controlled trial enrolled obese participants with metabolic dysfunction-associated steatohepatitis (MASH) and type 2 diabetes. After a 2-week screening period, eligible participants were randomly assigned in a 1:1 ratio to receive either linafexor (4 mg orally once daily) plus semaglutide (0.5 mg subcutaneously once weekly) or placebo plus semaglutide (0.5 mg subcutaneously once weekly) for 16 weeks. The primary end point was assessed at week 16, followed by a 4-week off-treatment follow-up period to evaluate safety and potential rebound effects. All participants received standardized lifestyle counseling throughout the trial.

### Extended Data Fig. 2 | Primary Outcome According to Prespecified Subgroup

| Subgroup | N | ETD (95% CI)* | P value for interaction |
|---|---|---|---|
| Overall | 62 | -3.26 (-5.35 to -1.18) | |
| Age | | | 0.85 |
| <40 years | 38 | -5.59 (-8.18 to -3.00) | |
| ≥40 years | 24 | -1.36 (-5.18 to 2.47) | |
| Sex | | | 0.52 |
| Male | 43 | -3.50 (-6.15 to -0.86) | |
| Female | 19 | -2.77 (-6.18 to 0.64) | |
| Body mass index | | | 0.97 |
| ≤35 kg/m² | 38 | -2.01 (-4.83 to 0.80) | |
| >35 kg/m² | 24 | -5.03 (-7.95 to -2.12) | |
| Liver fat severity | | | 0.93 |
| Moderate or mild liver fat | 41 | -3.52 (-6.19 to -0.86) | |
| Severe liver fat | 21 | -2.51 (-6.13 to 1.11) | |
| Glycated hemoglobin | | | 0.08 |
| ≤8.5% | 30 | -0.92 (-3.73 to 1.90) | |
| >8.5% | 32 | -5.42 (-8.38 to -2.46) | |
| Alanine aminotransferase | | | 0.24 |
| <40 IU/L | 37 | -4.27 (-6.80 to -1.74) | |
| ≥40 IU/L | 25 | -2.75 (-6.06 to 0.56) | |

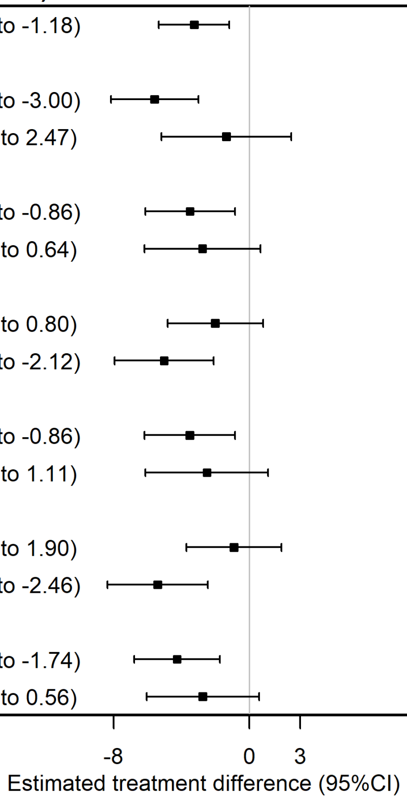


Forest plot demonstrating subgroup analyses for the primary outcome (difference in percentage change in body weight). Prespecified subgroups include age, sex, baseline body mass index, liver fat severity, glycated hemoglobin levels, and alanine aminotransferase levels. Vertical line indicates standard mean difference of 0 (no difference between groups). Square represents point estimates for the treatment effect (difference between linafexor + semaglutide and placebo + semaglutide), and horizontal lines indicate 95% confidence intervals. P values indicate subgroup interactions (obtained using mixed-effects models).

**Extended Data Fig. 3 | Plasma Concentration-Time Profiles of Linafexor at Week 16**

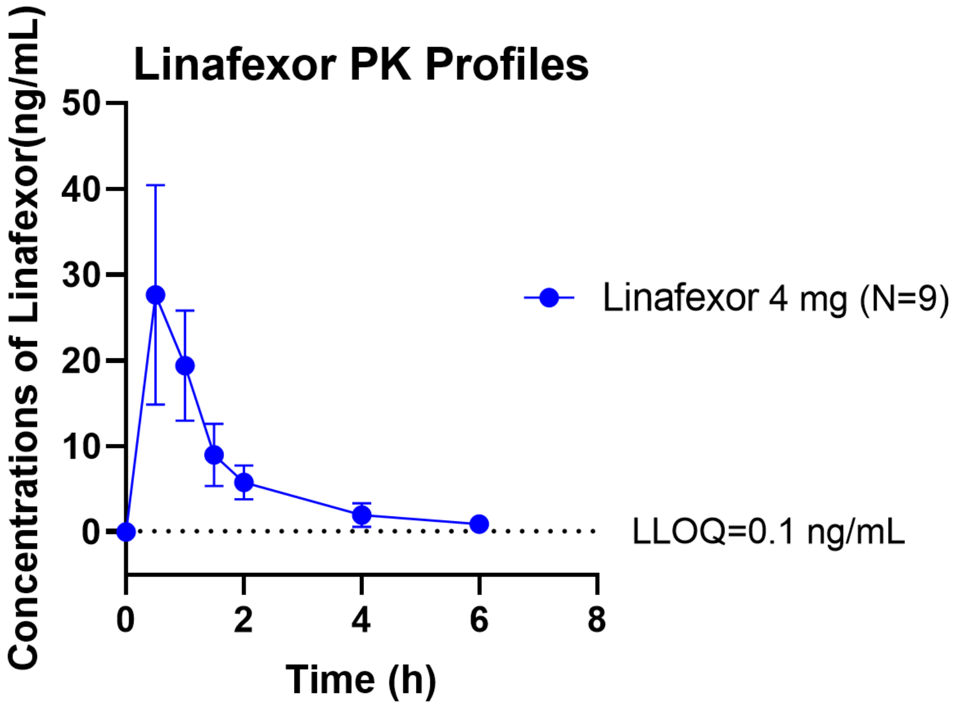


Pharmacokinetic (PK) concentration-time profiles of linafexor in 9 participants in the linafexor plus semaglutide group who underwent PK assessments at Week 16. Blood samples were collected at the following time points: pre-dose (within 30 minutes before administration), and post-dose at 30 minutes (±5 min), 1 hour (±10 min), 1.5 hours (±10 min), 2 hours (±10 min), 4 hours (±10 min), and 6 hours (±10 min). All PK values in the Placebo plus semaglutide group were below the Lower Limit of Quantification (LLOQ = 0.1 ng/mL).

**Extended Data Fig. 4 | Time to Onset of First Gastrointestinal Adverse Event**

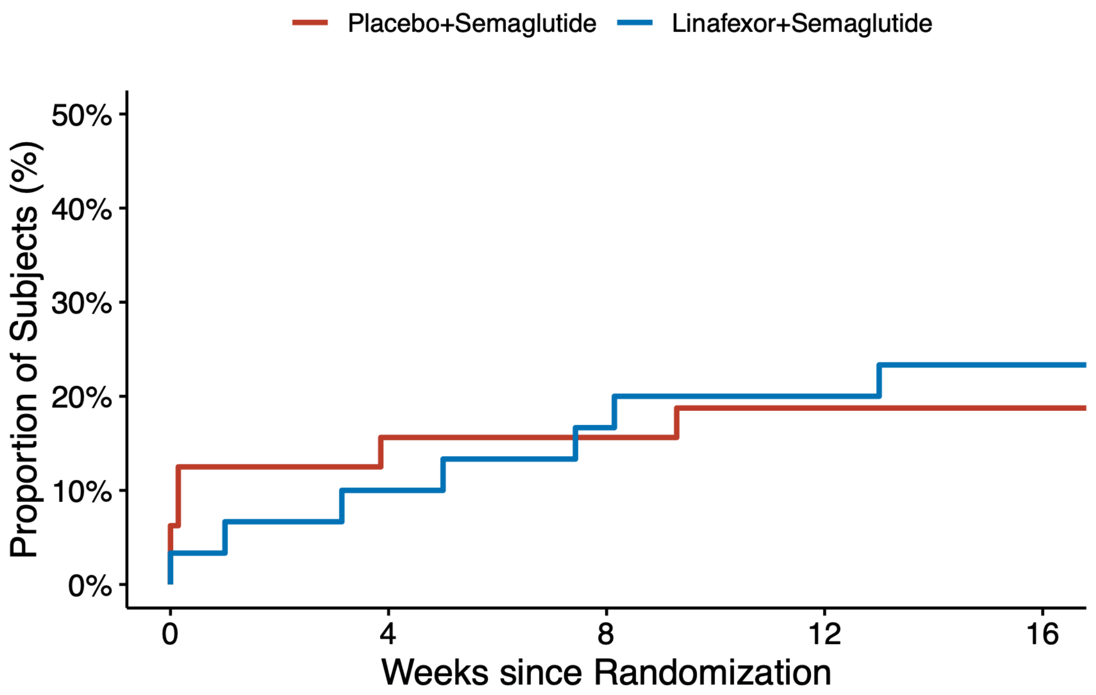


Data are on-treatment observation period data with Safety Analysis Set. All gastrointestinal adverse events were mild and moderate.

### Extended Data Table 1 | Primary and Key Secondary End Points at Week 16 (Treatment Policy Estimand)*

| Variable | Linafexor+Semaglutide N=30† | Placebo+Semaglutide N=32† | Treatment Difference (95% CI) ‡ | P value |
|---|---|---|---|---|
| **Primary end point** | | | | |
| **Percentage change in Body weight - %** | -6.17 (-7.64, -4.70) | -2.91 (-4.48, -1.33) | -3.32 (-5.50, -1.14) | 0.004 |
| **Secondary end points** | | | | |
| ≥5% reduction in body weight - %§ | 53.3 (34.7, 71.1) | 28.1 (13.7, 46.7) | 2.92 (1.02, 8.37) | 0.046 |
| Change in body mass index - kg/m$^2$ | -2.10 (-2.60, -1.59) | -0.96 (-1.51, -0.40) | -1.15 (-1.89, -0.42) | 0.002 |
| Change in waist circumference - cm | -5.78 (-7.80, -3.76) | -4.87 (-6.69, -3.04) | -0.75 (-2.66, 1.16) | 0.44 |
| Change in fat mass - kg¶ | -3.66 (-5.60, -1.72) | -1.58 (-3.21, 0.04) | -1.53 (-3.31, 0.26) | 0.092 |
| Change in fat percentage - %¶ | -2.32 (-3.61, -1.03) | -1.36 (-2.42, -0.30) | -0.70 (-1.87, 0.48) | 0.24 |
| Change in skeletal muscle mass - kg¶ | -0.31 (-0.95, 0.33) | 0.19 (-0.31, 0.68) | -0.37 (-0.94, 0.20) | 0.20 |
| Change in visceral adipose tissue area - cm$^2$¶ | -35.60 (-46.32, -24.88) | -13.95 (-24.75, -3.14) | -13.85 (-24.52, -3.17) | 0.035 |
| Change in subcutaneous adipose tissue area - cm$^2$¶ | -18.99 (-33.63, -4.36) | -13.00 (-26.61, 0.61) | -2.92 (-16.65, 10.81) | 0.67 |
| Change in glycated hemoglobin - % | -1.98 (-2.60, -1.35) | -2.28 (-2.73, -1.83) | 0.55 (-4.71, 5.80) | 0.81 |
| Change in fasting plasma glucose - mg/dL | -42.15 (-53.57, -30.73) | -52.91 (-67.64, -38.19) | 3.85 (-6.49, 14.19) | 0.46 |
| Change in 2h post-prandial plasma glucose - mg/dL | -94.79 (-125.13, -64.46) | -96.68 (-118.89, -74.48) | -1.31 (-22.10, 19.48) | 0.90 |
| Percentage change in lipid levels | | | | |
| Total cholesterol | 1.46 (-7.07, 9.98) | -5.37 (-10.80, 0.06) | 2.33 (-3.52, 8.17) | 0.43 |
| Triglycerides | -18.69 (-35.09, -2.28) | -25.67 (-38.41, -12.92) | 1.55 (-11.72, 14.82) | 0.82 |
| High-density lipoprotein cholesterol | -1.07 (-6.06, 3.92) | 0.77 (-4.23, 5.77) | -0.88 (-5.37, 3.61) | 0.70 |
| Low-density lipoprotein cholesterol | 8.68 (-2.20, 19.55) | 11.41 (-12.71, 35.53) | -6.11 (-20.79, 8.58) | 0.41 |
| Non-high-density lipoprotein cholesterol | 3.38 (-7.44, 14.20) | -6.03 (-12.63, 0.57) | 3.20 (-4.09, 10.48) | 0.38 |
| Percentage change in liver enzyme levels | | | | |
| Alanine aminotransferase | -45.31 (-58.08, -32.55) | -23.79 (-41.41, -6.18) | -12.83 (-24.76, -0.91) | 0.035 |

| | | | | |
|---|---|---|---|---|
| Aspartate aminotransferase | -32.19 (-45.30, -19.08) | -24.98 (-39.04, -10.91) | 1.90 (-7.34, 11.13) | 0.68 |
| γ-Glutamyl transferase | -52.84 (-61.62, -44.06) | -22.94 (-32.31, -13.57) | -20.01 (-28.18, -11.85) | <0.001 |
| Alkaline phosphatase | -2.34 (-9.17, 4.48) | -18.86 (-23.92, -13.80) | 11.06 (5.52, 16.59) | <0.001 |
| Percentage change in liver stiffness‖ | -2.71 (-4.56, -0.86) | -1.72 (-3.85, 0.41) | -0.99 (-3.80, 1.80) | 0.67 |
| Percentage change in liver fat content** | -45.54 (-57.16, -33.92) | -35.79 (-47.08, -24.50) | -7.77 (-18.86, 3.32) | 0.17 |

* Shown are estimated data for the treatment policy estimand, which was used to assess the treatment effect in the full analysis population regardless intercurrent events. For the primary end point and other continuous end points were assessed with the use of analysis of covariance, and categorical end points were analyzed with the use of logistic regression, including treatment group, randomization stratification factor, and baseline value of the endpoint as covariates. Missing data was handled using multiple imputation. Since no intercurrent events (i.e., trial treatment discontinuation or rescue medication initiation) occurred during the study, the treatment policy estimand values for non-repeated endpoints were identical to the trial product estimand estimates presented in Table 2.

† Data are the mean change from baseline to Week 16, with 95% confidence intervals (CI).

‡ Data are estimated mean difference between two treatment groups.

§ Data are the observed percentages of participants at week 16, and the difference is the estimated odds ratios.

¶ Body composition including fat mass, fat percentage, and skeletal muscle mass was measured by means of bioelectrical impedance analysis (BIA). Visceral adipose tissue (VAT) area and subcutaneous adipose tissue (SAT) area were measured by magnetic resonance imaging (MRI) acquired measurement.

|| Liver stiffness was assessed by vibration-controlled transient elastography (VCTE).

** Liver fat content was assessed by means of the MRI proton density fat fraction (MRI-PDFF).

### Extended Data Table 2 | Primary and Key Secondary End Points at Week 16 (Per-Protocol Set)*

| Variable | Linafexor+Semaglutide N=29† | Placebo+Semaglutide N=31† | Treatment Difference (95% CI) ‡ | P value |
|---|---|---|---|---|
| **Primary end point** | | | | |
| **Percentage change in Body weight - %** | -6.32 (-7.81, -4.83) | -2.58 (-4.08, -1.09) | -3.67 (-5.72,-1.63) | 0.001 |
| **Secondary end points** | | | | |
| ≥5% reduction in body weight - %§ | 55.2 (36.03, -73.0) | 25.8 (13.7, 46.7) | 3.54 (1.19, 10.5) | 0.023 |
| Change in body mass index - $kg/m^2$ | -2.15 (-2.66, -1.64) | -0.85 (-1.38, -0.32) | -1.29 (-1.97,-0.61) | <0.001 |
| Change in waist circumference - cm | -5.98 (-8.03, -3.93) | -4.97 (-6.84, -3.10) | -0.67 (-2.62, 1.27) | 0.49 |
| Change in fat mass - kg¶ | -3.79 (-5.78, -1.79) | -1.24 (-2.76, 0.27) | -1.77 (-3.51, -0.03) | 0.047 |
| Change in fat percentage - %¶ | -2.40 (-3.72, -1.07) | -1.14 (-2.13, -0.15) | -0.85 (-1.99, 0.29) | 0.14 |
| Change in skeletal muscle mass - kg¶ | -0.32 (-0.99, 0.34) | 0.22 (-0.28, 0.73) | -0.39 (-0.96, 0.19) | 0.18 |
| Change in visceral adipose tissue area - $cm^2$¶ | -35.78 (-46.88, -24.67) | -12.60 (-23.41, -1.79) | -14.96 (-25.84, -4.08) | 0.13 |
| Change in subcutaneous adipose tissue area - $cm^2$¶ | -20.46 (-35.31, -5.61) | -12.99 (-27.06, 1.09) | -4.00 (-18.10, 10.09) | 0.57 |
| Change in glycated hemoglobin - % | -2.03 (-2.66, -1.39) | -2.27 (-2.74, -1.80) | 0.04 (-0.40, 0.49) | 0.85 |
| Change in fasting plasma glucose - mg/dL | -42.05 (-53.89, -30.22) | -52.80 (-68.03, -37.58) | 3.78 (-6.75, 14.30) | 0.47 |
| Change in 2h post-prandial plasma glucose - mg/dL | -98.06 (-128.73, -67.39) | -97.73 (-120.59, -74.88) | -0.61 (-21.71, 20.49) | 0.95 |
| Percentage change in lipid levels - mmol/L | | | | |
| Total cholesterol | 0.48 (-8.12, 9.08) | -4.76 (-10.23, 0.71) | 1.29 (-4.53, 7.10) | 0.66 |
| Triglycerides | -24.16 (-36.59, -11.74) | -25.15 (-38.28, -12.01) | -3.30 (-14.13, 7.53) | 0.54 |
| High-density lipoprotein cholesterol | -1.14 (-6.31, 4.03) | 1.35 (-3.68, 6.38) | -1.12 (-5.76, 3.52) | 0.63 |
| Low-density lipoprotein cholesterol | 8.36 (-2.89, 19.62) | 12.52 (-12.31, 37.36) | -7.05 (-22.16, 8.06) | 0.35 |
| Non-high-density lipoprotein cholesterol | 2.23 (-8.72, 13.17) | -5.37 (-12.05, 1.31) | 1.84 (-5.41, 9.10) | 0.61 |
| Percentage change in liver enzyme levels - U/liter | | | | |
| Alanine aminotransferase | -47.23 (-59.82, -34.63) | -22.35 (-40.31, -4.39) | -14.95 (-26.90, -3.00) | 0.015 |

| | | | | |
|---|---|---|---|---|
| Aspartate aminotransferase | -35.34 (-47.18, -23.50) | -24.13 (-38.57, -9.69) | -0.45 (-9.43, 8.52) | 0.92 |
| γ-Glutamyl transferase | -54.70 (-62.90, -46.50) | -22.31 (-31.91, -12.71) | -21.95 (-29.77, -14.12) | <0.001 |
| Alkaline phosphatase | -1.92 (-8.94, 5.09) | -19.79 (-24.64, -14.94) | 11.82 (6.19, 17.45) | <0.001 |
| Percentage change in liver stiffness‖ | -2.80 (-4.71, -0.90) | -1.68 (-3.88, 0.52) | -0.35 (-1.72, 1.02) | 0.61 |
| Percentage change in liver fat content** | -47.11 (-58.69, -35.53) | -34.48 (-45.83, -23.14) | -8.61 (-19.75, 2.54) | 0.13 |

* Shown are estimated data for the per-protocol set. Outcome measurements after the intercurrent event were treated as missing data. For the primary end point and other repeatedly measured weight loss-related outcomes, the mixed-effect model was used to estimate treatment difference between groups, which includes treatment group, visit time, and their interaction as fixed effects. Non-repeated measurement continuous end points were assessed with the use of analysis of covariance, including treatment group, randomization stratification factor, and baseline value of the endpoint as covariates. Categorical end points were analyzed with the use of logistic regression, with the same factors and covariate. Missing data was handled using multiple imputation approach.

† Data are the mean change from baseline to Week 16, with 95% confidence intervals (CI).

‡ Data are estimated mean difference between two treatment groups.

§ Data are the observed percentages of participants at week 16, and the difference is the estimated odds ratios.

¶ Body composition including fat mass, fat percentage, and skeletal muscle mass was measured by means of BIA. VAT area and SAT area were measured by MRI acquired measurement.

|| Liver stiffness was assessed by VCTE.

** Liver fat content was assessed by means of the MRI-PDFF.

**Extended Data Table 3 | Association Between Intervention and Changes in Liver fat content and Visceral Adipose Tissue Area**

| | Model 1 | | Model 2 | |
|---|---|---|---|---|
| | Difference (95% CI) | P value | Difference (95% CI) | P value |
| Liver fat content | -8.4 (-19.0, 2.2) | 0.12 | -11.5 (-21.5, -1.5) | 0.025 |
| Visceral adipose tissue area (MRI) | -15.2 ( -25.4, -5.0 ) | 0.004 | -16.8 ( -27.3, -6.3 ) | 0.002 |

Results are presented as differences (95% CI) between intervention groups. Liver fat content was measured using MRI-PDFF. VAT area was assessed using MRI acquired measurement.

Model 1: Multivariable linear regression, adjusted for age, sex, and baseline value.

Model 2: Multivariable linear regression, adjusted for age, sex, baseline value, body mass index, glycated hemoglobin, drink status, smoking status.

**Extended Data Table 4 | Differential bile acids before and after 16-week intervention in the Linafexor plus Semaglutide group**

| | Fasting state | | | | | Postprandial state | | | | |
|---|---|---|---|---|---|---|---|---|---|---|
| Metabolite | Baseline Median [IQR] | Post-treatment Median [IQR] | Fold Change | P value | FDR | Baseline Median [IQR] | Post-treatment Median [IQR] | Fold Change | P value | FDR |
| βCA | 8.44 [2.51,16.85] | 2.17 [0.89,5.32] | 0.256 | <0.001 | 0.004 | 10.23 [1.42,25.72] | 1.6 [0.98,3.30] | 0.156 | <0.001 | 0.001 |
| GCA | 115.74 [72.40,191.96] | 29.76 [21.03,71.50] | 0.257 | <0.001 | <0.001 | 117.67 [63.39,211.41] | 28.69 [9.14,65.28] | 0.244 | <0.001 | <0.001 |
| βDCA | 45.3 [24.50,124.40] | 13.58 [6.31,55.30] | 0.300 | <0.001 | 0.001 | 47.24 [17.95,95.39] | 10.61 [4.25,33.35] | 0.225 | <0.001 | <0.001 |
| GHCA | 3.8 [2.46,9.26] | 1.37 [0.74,2.85] | 0.361 | <0.001 | 0.004 | 4.29 [2.51,6.98] | 1.03 [0.82,2.66] | 0.241 | <0.001 | 0.001 |
| GCDCA | 422.03 [267.37,793.87] | 182.33 [93.54,493.20] | 0.432 | 0.003 | 0.016 | 398.39 [189.11,671.26] | 149.4 [54.78,300.39] | 0.375 | <0.001 | 0.002 |
| TCA | 25.85 [11.44,39.70] | 11.26 [5.01,20.94] | 0.435 | 0.010 | 0.035 | 19.68 [12.60,47.39] | 8.93 [4.13,16.56] | 0.454 | 0.002 | 0.008 |
| βUDCA | 198.57 [71.16,627.91] | 88.58 [26.09,299.25] | 0.446 | 0.041 | 0.110 | 227.83 [78.08,481.26] | 57.98 [21.71,281.77] | 0.254 | 0.003 | 0.010 |
| GUDCA | 92.52 [26.03,198.15] | 41.49 [15.51,109.66] | 0.448 | 0.029 | 0.085 | 49.98 [22.45,140.00] | 20.79 [8.19,79.33] | 0.416 | 0.006 | 0.016 |
| 6-KetoLCA | 2.67 [1.85,3.11] | 1.55 [1.02,2.30] | 0.579 | 0.002 | 0.014 | 2.25 [1.75,3.20] | 1.24 [0.57,1.71] | 0.553 | <0.001 | 0.001 |
| NorDCA | 0.26 [0.03,0.66] | 1.59 [0.36,2.25] | 6.144 | 0.002 | 0.013 | 0.42 [0.14,0.72] | 0.82 [0.34,2.05] | 1.946 | 0.036 | 0.083 |

| | | | | | | | | | | |
|---|---|---|---|---|---|---|---|---|---|---|
| LCA-3S | 1.43 [0.33,4.91] | 5.97 [1.54,9.97] | 4.187 | 0.013 | 0.041 | NS | NS | NS | NS | NS |
| TLCA | 0.21 [0.09,0.60] | 0.6 [0.27,1.44] | 2.848 | 0.009 | 0.033 | 0.34 [0.09,0.78] | 0.96 [0.19,1.56] | 2.830 | 0.026 | 0.065 |
| CA | 47.03 [27.44,96.93] | 132.76 [18.62,433.71] | 2.823 | 0.005 | 0.021 | NS | NS | NS | NS | NS |
| 12-DHCA | NS | NS | NS | NS | NS | 5.03 [1.99,8.60] | 2.02 [1.10,3.28] | 0.402 | 0.004 | 0.014 |
| 3-DHCA | NS | NS | NS | NS | NS | 5.63 [3.80,8.07] | 2.46 [0.90,4.76] | 0.437 | <0.001 | 0.002 |
| CDCA-3Glu | NS | NS | NS | NS | NS | 16.1 [10.90,43.47] | 11.99 [7.01,20.61] | 0.745 | 0.002 | 0.007 |
| UCA | NS | NS | NS | NS | NS | 2.2 [0.67,6.23] | 1.64 [0.91,3.20] | 0.745 | 0.014 | 0.037 |

Significantly altered bile acids after 16-week intervention in the Linafexor plus Semaglutide group under fasting and postprandial states. Bile acid concentrations are expressed in nmol/L. Data are presented as median (interquartile range [IQR]) and fold change. P-values were calculated using the Wilcoxon signed-rank test, and adjusted for false discovery rate (FDR) using the Benjamini-Hochberg method.

βCA, 3β-Cholic Acid; GCA, Glycocholic Acid; βDCA, 3β-Deoxycholic Acid; GHCA, Glycohyocholic Acid; GCDCA, Glycochenodeoxycholic Acid; TCA, Taurocholic Acid; βUDCA, Isoursodeoxycholic Acid; GUDCA, Glycoursodeoxycholic Acid; 6-KetoLCA, 6-Ketolithocholic Acid; NorDCA, Nordeoxycholic Acid; LCA-3S, Lithocholic Acid 3-Sulfate; TLCA, Taurolithocholic Acid; CA, Cholic Acid; 12-DHCA, 12-Dehydrocholic Acid; 3-DHCA, 3-Oxocholic Acid; CDCA-3Glu, Chenodeoxycholic Acid-3-β-D-Glucuronide; UCA, Ursocholic Acid;

**Extended Data Table 5 | Differential bile acids before and after 16-week intervention in the Placebo plus Semaglutide group**

| | Fasting state | | | | | Postprandial state | | | | |
|---|---|---|---|---|---|---|---|---|---|---|
| Metabolite | Baseline Median [IQR] | Post-treatment Median [IQR] | Fold Change | P value | FDR | Baseline Median [IQR] | Post-treatment Median [IQR] | Fold Change | P value | FDR |
| βDCA | 52.84 [34.71,97.19] | 21.67 [11.06,49.16] | 0.410 | 0.019 | 0.068 | NS | NS | NS | NS | NS |
| GCDCA | 769.71 [279.10,1103.24] | 333.37 [167.05,613.29] | 0.433 | 0.001 | 0.006 | NS | NS | NS | NS | NS |
| TUDCA | 2.41 [0.77,6.19] | 1.06 [0.70,2.36] | 0.438 | 0.017 | 0.068 | NS | NS | NS | NS | NS |
| TCA | 38.22 [20.50,69.45] | 18.63 [7.36,29.39] | 0.487 | <0.001 | 0.006 | NS | NS | NS | NS | NS |
| GCA | 207.2 [71.58,363.17] | 105.61 [34.63,164.94] | 0.510 | <0.001 | 0.006 | NS | NS | NS | NS | NS |
| TCDCA | 75.15 [31.47,118.61] | 38.31 [18.28,61.57] | 0.510 | <0.001 | 0.006 | NS | NS | NS | NS | NS |
| 6-KetoLCA | 2.1 [1.59,3.78] | 1.36 [1.02,2.06] | 0.649 | 0.001 | 0.009 | NS | NS | NS | NS | NS |
| GUDCA | 73.94 [42.55,178.67] | 48.63 [29.25,118.72] | 0.658 | 0.041 | 0.115 | NS | NS | NS | NS | NS |
| CA | 44.08 [20.15,91.11] | 160.91 [36.26,384.54] | 3.650 | 0.007 | 0.032 | 30.88 [15.06,67.98] | 52.08 [26.04,171.02] | 1.687 | 0.027 | 0.223 |
| DCA-3S | 0.99 [0.90,2.42] | 2.98 [0.96,5.42] | 3.028 | 0.036 | 0.114 | 0.98 [0.42,1.68] | 2.05 [0.96,4.79] | 2.078 | 0.002 | 0.084 |

| | | | | | | | | | | |
|---|---|---|---|---|---|---|---|---|---|---|
| 7-DHCA | 2.55 [0.85,5.72] | 7.57 [1.65,12.05] | 2.972 | 0.002 | 0.010 | NS | NS | NS | NS | NS |
| LCA-3S | 3.25 [1.10,8.78] | 7.39 [2.05,13.37] | 2.277 | 0.043 | 0.115 | 2.25 [0.92,5.17] | 4.69 [1.60,9.29] | 2.084 | 0.005 | 0.088 |
| HCA | 4.97 [1.60,8.20] | 9.67 [5.78,15.01] | 1.946 | 0.002 | 0.009 | 2.93 [1.48,7.82] | 7.48 [2.67,9.93] | 2.550 | 0.031 | 0.223 |
| HDCA | NS | NS | NS | NS | NS | 0.55 [0.14,1.68] | 0.99 [0.39,2.11] | 1.805 | 0.032 | 0.223 |

Significantly altered bile acids after 16-week intervention in the Placebo plus Semaglutide group under fasting and postprandial states. Bile acid concentrations are expressed in nmol/L. Data are presented as median (interquartile range [IQR]) and fold change. P-values were calculated using the Wilcoxon signed-rank test, and adjusted for false discovery rate (FDR) using the Benjamini-Hochberg method.

βDCA, 3β-Deoxycholic Acid; GCDCA, Glycochenodeoxycholic Acid; TUDCA, Tauroursodeoxycholic Acid; TCA, Taurocholic Acid; GCA, Glycocholic Acid; TCDCA, Taurochenodeoxycholic Acid; 6-KetoLCA, 6-Ketolithocholic Acid; GUDCA, Glycoursodeoxycholic Acid; CA, Cholic Acid; DCA-3S, 3-Sulfodeoxycholic Acid; 7-DHCA, 7-Dehydrocholic Acid; LCA-3S, Lithocholic Acid 3-Sulfate; HCA, Hyocholic Acid; HDCA, Hyodeoxycholic Acid;
